# SARS-CoV-2 induces human endogenous retrovirus type W envelope protein expression in blood lymphocytes and in tissues of COVID-19 patients

**DOI:** 10.1101/2022.01.18.21266111

**Authors:** Benjamin Charvet, Joanna Brunel, Justine Pierquin, Mathieu Iampietro, Didier Decimo, Nelly Queruel, Alexandre Lucas, María del Mar Encabo-Berzosa, Izaskun Arenaz, Tania Perez Marmolejo, Francina Valezka Bolaños Morales, Arturo Ivan Gonzalez Gonzalez, Armando Castorena Maldonado, César Luna Rivero, Cyrille Mathieu, Patrick Küry, Jose Flores-Rivera, Santiago Avila Rios, Gonzalo Salgado Montes de Oca, Jon Schoorlemmer, Branka Horvat, Hervé Perron

**Affiliations:** GeNeuro Innovation, Lyon, France.; CIRI, International Center for Infectiology Research, INSERM U1111, CNRS UMR5308, Université de Lyon, Université Claude Bernard Lyon 1, Ecole Normale Supérieure de Lyon, France; We-Met platform, I2MC/Inserm/Université Paul Sabatier UMR1297, Toulouse, France; Biobanco del Sistema de Salud de Aragón, Instituto Aragonés de Ciencias de la Salud (IACS), Zaragoza, Spain; Instituto Nacional de Enfermedades Respiratorias Ismael Cosio Villegas, México Ciudad, México.; Department of Neurology, Medical Faculty, Heinrich-Heine-University, Dusseldorf, Germany; Department of Neurology, National Institute of Neurology and Neurosurgery, Mexico City, Mexico; Centro de investigación en Enfermedades Infecciosas, Instituto Nacional de Enfermedades Respiratorias Ismael Cosío Villegas, México Ciudad, México; ARAID Fundación; Instituto Aragonés de Ciencias de la Salud (IACS); Grupo B46_20R de la DGA and GIIS-028 del IISA; all Zaragoza, Spain; GeNeuro, Plan les Ouates, Geneva, Switzerland

## Abstract

Patients with COVID-19 may develop abnormal inflammatory response and lymphopenia, followed in some cases by delayed-onset syndromes, often long-lasting after the initial SARS-CoV-2 infection. As viral infections may activate human endogenous retroviral elements (HERV), we studied the effect of SARS-CoV-2 on HERV-W and HERV-K envelope (ENV) expression, known to be involved in immunological and neurological pathogenesis of human diseases. Our results have showed that the exposure to SARS-CoV-2 virus activates early HERV-W and K transcription but only HERV-W ENV protein expression, in an infection- and ACE2-independent way within peripheral blood mononuclear cell cultures from one-third of healthy donors. Moreover, HERV-W ENV protein was significantly increased in serum and plasma of COVID-19 patients, correlating with its expression in CD3^+^ lymphocytes and with disease severity. Finally, HERV-W ENV was found expressed in post-mortem tissues of lungs, heart, brain olfactory bulb and nasal mucosa from acute COVID-19 patients in cell-types relevant for COVID-19-associated pathogenesis within affected organs, but different from those expressing of SARS-CoV-2 antigens. Altogether, the present study revealed that SARS-CoV-2 can induce HERV-W ENV expression in cells from individuals with symptomatic and severe COVID-19. Our data suggest that HERV-W ENV is likely to be involved in pathogenic features underlying symptoms of acute and post-acute COVID. It highlights the importance to further understand patients’ genetic susceptibility to HERV-W activation and the relevance of this pathogenic element as a prognostic marker and a therapeutic target in COVID-19 associated syndromes.

**Graphical abstract:** 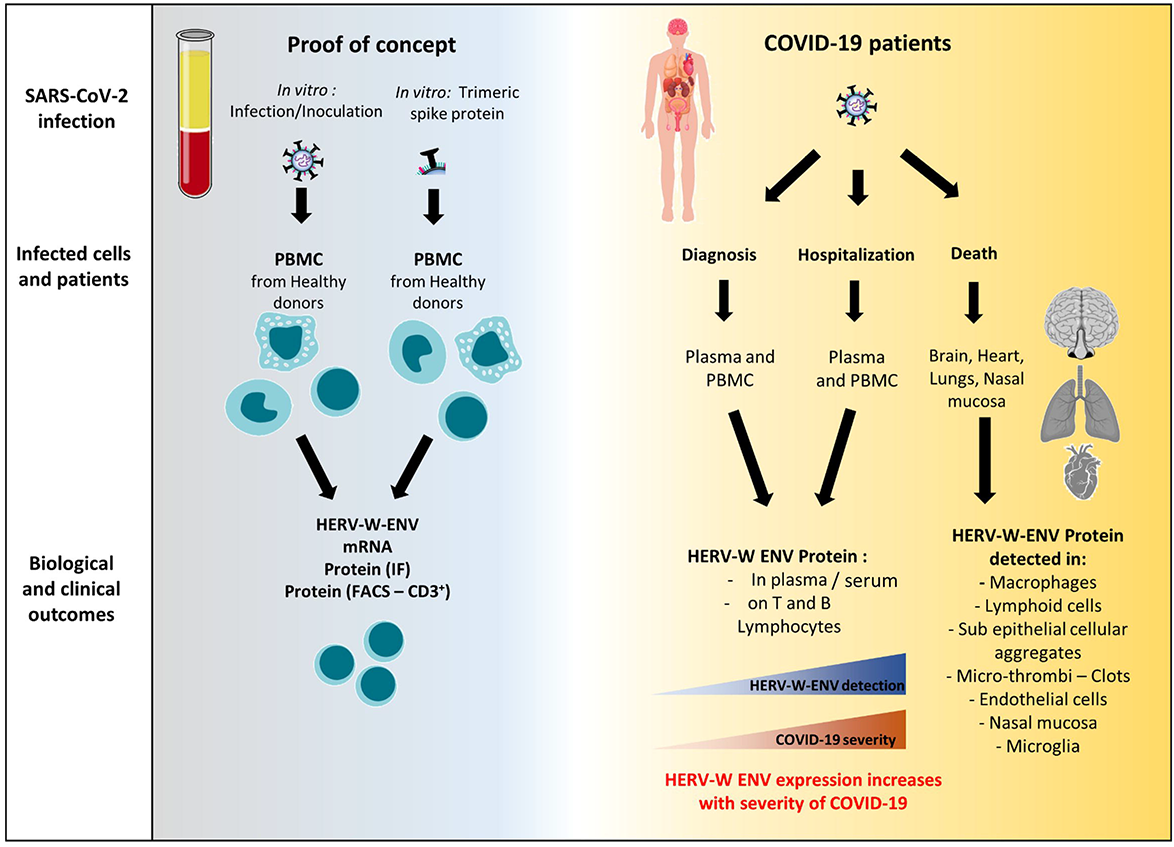

## Introduction

The human “coronavirus disease 2019” (COVID-19) caused by the severe acute respiratory syndrome coronavirus 2 (SARS-CoV-2) is associated with high morbidity and mortality [1–6]. It has generated a worldwide pandemic, the evolution of which is still uncertain due to the emergence of variants [7–11]. Hospitals worldwide have been facing the increasing demand of patients requiring oxygen supply, ventilators, and intensive care. Vaccination helps to progressively decrease the number of severe primary infections but numerous long-lasting and/or post-infectious symptoms or syndromes are commonly observed among patients who had COVID-19, from benign to severe forms [12–18]. Moreover, beyond a dominant respiratory tract tropism, extra-pulmonary COVID-19 forms are more frequent and diverse than expected [19].

The dysregulation of innate and adaptive immunity has been recognized to play a critical role in the clinical outcome of COVID-19 patients. Severe evolution of COVID-19 is thought to be driven by hyperactivated innate immunity [20–22], in addition to adaptive immune defects resulting in lymphopenia and neutrophils/lymphocytes imbalance [23, 24]. A deficient interferon response has also been shown to favor or result from SARS-CoV-2 infection [25, 26]. Such multifaceted immunological dysregulations are thus underlying hyper-immune reactions such as the “cytokine storm” syndrome, the multisystem inflammatory syndrome in children (MIS-C), the dysregulation of coagulation, as well as neurological and various other manifestations [27–29]. The present COVID-19 pandemic has thus raised many questions about the pathophysiological mechanisms explaining the many symptoms or syndromes associated with SARS-CoV-2 infection.

Certain infectious agents have been shown to activate pathological processes via receptor-coupled signaling pathways, by impairing the epigenetic control and/or by directly activating endogenous retroviral elements (HERVs) present in the human genome [30, 31]. In particular, the resulting production of endogenous proteins of retroviral origin with pathogenic effects may generate clinical symptoms corresponding to the organ, tissue or cells in which they are expressed, according to the specific tropism of the triggering infectious agent [32–40]. HERVs represent about 8% of human chromosomal sequences and comprise about 22 families independently acquired during evolution from exogenous retroviruses via an infection of germ line cells [41, 42]. Abnormal expression may then become self-sustained, thus creating lifelong chronic expression from host’s genome copies in affected tissues [43], e.g., with cytokine-mediated feedback loops [44] or, possibly, mediated by their own envelope proteins [45]. Such a sustained expression has been shown to be involved in brain lesions with lifelong expansion in patients with multiple sclerosis (MS) [43, 46–48]. Different HERV envelope proteins were shown to exert major immunopathogenic [49–55] and/or neuropathogenic [46, 51–58] effects *in vitro* and *in vivo*, associated with pathognomonic features of human diseases. We therefore studied whether SARS-CoV-2 could activate HERV copies considered as ‘dormant enemies within’ [59]. We focused on HERV families already shown to be involved in the pathogenesis of human diseases, HERV-W and HERV-K [57], to evaluate their potential association with COVID-19 and associated syndromes. This question became critical after a recent study has revealed the significant expression of HERV-W envelope protein (ENV) in lymphoid cells from COVID-19 patients, correlating with disease outcome and markers of lymphocyte exhaustion or senescence [60].

In the present study, we initially addressed the potential role of SARS-CoV-2 in directly triggering the activation of a pathogenic HERV protein expression as reported with other viruses in, e.g., MS and in type 1 diabetes [61–63]. We further analyzed its expression in white blood cells and its possible detection in plasma of patients with COVID-19 presenting various clinical forms at early and late time-points. We finally examined this HERV expression in affected tissues from COVID-19 post-mortem samples. Also, since a major concern beyond the initial COVID-19 infectious phase is foreseen to result from the occurrence of long lasting symptoms with more-or-less delayed onset and often involves neurological impairment [17, 64–66], we examined SARS-CoV-2 and HERV-W expression in brain parenchyma. For comparison with dominantly or frequently affected organs, we also examined these antigens expression in lung and cardiac tissues.

Our results showed that (i) SARS-CoV-2 can activate the production of HERV-W ENV in cultured blood mononuclear cells from a sub-group of healthy donors, (ii) HERV-W ENV is expressed on T-lymphocytes from COVID-19 patients, (iii) HERV-W ENV antigen is detected in all tested plasma or sera samples from severe cases in intensive care unit, but only in about 20% of PCR positive cases after early diagnosis, (iv) the level of HERV-W ENV antigenemia increases with disease severity and (iv) HERV-W ENV expression is observed by immunohistochemistry in cell-types relevant for COVID-19 associated pathogenesis within affected organs and particularly in brain microglia.

## Materials and Methods

### A. In vitro series

#### A1. Cells

Peripheral blood mononuclear cells (PBMC) from healthy donors were obtained from the “Etablissement Français du Sang” (EFS) of Lyon (France). PBMC were isolated by Ficoll separation (Ficoll-Plaque PLUS) (GE Healthcare, 17-1440-02) from blood samples and cultured in RPMI-1640 medium (Gibco, 61870-010) completed with 5% of decomplemented Human AB serum (Sigma, H4522). Healthy donors signed a written Informed Consent Form, documented at the EFS, allowing the commercial use of their blood and blood components for medical research after definite anonymization.

Vero E6 cells were grown in DMEM Glutamax (Thermo) supplemented with 10% fetal bovine serum (FBS), glutamine and antibiotics (100 U/mL of penicillin and 100 µg/mL of streptomycin) in 5% CO2 incubators at 37°C and were tested negative for mycoplasma spp.

#### A2. SARS-CoV-2 serology of blood donors

SARS-CoV-2 serology of blood donors was determined on plasma diluted 10 times using Simple Western technology, an automated capillary-based size sorting and immunolabeling system (ProteinSimple^TM^). The SARS-CoV-2 Multi-Antigen Serology Module (SA-001) was used with Wes device and all procedures were performed according to manufacturer’s protocol. Wes device was associated with Compass software for device settings and raw data recording (ProteinSimple/Biotechne).

#### A3. Exposure to wild-type SARS-CoV-2 or recombinant active trimer Spike SARS-CoV-2

SARS-CoV-2 strain (BetaCoV/France/IDF0571/2020, GISAID Accession ID: EPI_ISL_411218) was cultured on Vero E6 cell line (ATCC®CRL1586™) for virus production. All experiments were performed with freshly prepared PBMC of independent donors. PBMC and Vero cells were infected with SARS-CoV-2 at a multiplicity of infection (MOI) 0.1. PBMC inoculation with SARS-CoV-2 infectious virus at 0.1 MOI was performed in RPMI-1640 medium (Gibco, 61870-010) completed with 2% of heat inactivated Human AB serum (Sigma, H4522). Infection of Vero cells was performed in DMEM medium (DMEM, Gibco^TM^) completed with 2% of heat inactivated FCS. 2 h later, the concentrations of AB-human serum or FCS were increased to 10%. Treatment with recombinant non-stabilized trimer Spike SARS-CoV-2 (ACROBiosystems, USA; Ref. SPN-C52H8) was performed at 0.5 and 2.5 µg/mL.

#### A4. Immunofluorescence

Cells in suspension were pelleted by centrifugation and deposited on 3 well epoxy microscope slides (Thermo Scientific, 30-12A-BLACK-CE24) while adherent cells were manipulated directly in 48 wells plates. Suspension and adherent cells received the same following steps. Cells were fixed in paraformaldehyde 4% during 15 minutes at RT. Cells were washed tree times in 1X PBS and permeabilized 15 min in 0.2% Tween20, 1X PBS. Saturation was performed using 2.5% horse serum, 0.2% Tween20, 1 X PBS, during 30 minutes at room temperature before incubation with a mix of primary antibodies during 1 hour or overnight: 3 µg/mL of anti-HERV-W ENV (GeNeuro, GN_mAb_Env01, murine antibody) or 10 µg/mL anti-HERV-K ENV (GeNeuro, GN_mAb_Env-K01, murine antibody), together with either anti-N SARS-CoV-2 diluted 1/500 (SinoBiological, 40143-T62, rabbit antibody) or anti-S SARS-CoV-2 diluted 1/500 (SinoBiological, 40590-T62, rabbit antibody). Antibody solutions were prepared in the previously described saturation buffer. After three washes in PBS 1X, cells were incubated during 1 hour with secondary antibodies mix containing 1µg/mL goat anti-mouse Alexa Fluor 488 (ThermoFisher Scientific, A11029), 1 µg/mL donkey anti-rabbit Alexa Fluor 647 (ThermoFisher Scientific, A31573) and DAPI 1/2000 (Sigma, D9542) diluted in the previously described saturation buffer. Finally, cells were washed three times in 1X PBS and mounted using the Fluoromount-G mounting medium (Southern Biotech, 0100-01). Pictures were acquired on NIKON Eclipse TS2R microscope and analyzed on ImageJ software.

#### A5. Cytofluorometry

At 24 or 72 h post infection, cells were pelleted by centrifugation. PBMC were incubated with FcR Blocking Reagent according to manufacturer’s protocol (Miltenyi Biotec, 130-113-199) and stained with 1/10^e^ CD3-PE (BD Biosciences, 552127). Following cell surface staining, cells were fixed and permeabilized using Cytofix/Cytoperm kit (BD Biosciences, 554714) according to manufacturer’s instructions. Then, PBMC were stained with 10 µg/mL of an anti-HERV-W ENV-FITC (GeNeuro, GN_mAb_Env01-FITC) or mouse IgG1 FITC-labeled isotype control (Miltenyi Biotec, 130-113-199). Stained cells were acquired on a BD LSR Fortessa, fluorochrome emissions from the pool of antibodies were compensated using OneComp Beads (Invitrogen, 01-1111-42) and data analyzed with FlowJo software (v.10).

#### A6. Quantitative RT PCR (RT-qPCR)

RT-qPCR was performed using specific primers for HERV-W and HERV-K envelope genes, as already validated in patients with HERV-associated diseases [56, 62, 67], using *B2M* mRNA as a suitable reporter gene for PBMC [68].

For *in vitro* analyses, cells were harvested at several time points after exposure to SARS-CoV-2 virus or protein, and total RNA extracted. For blood samples, freshly isolated PBMC were collected to similarly extract RNA.

200 ng of DNase-treated RNA were reverse-transcribed into cDNA using iScript cDNA Synthesis Kit (Bio-Rad, 1708891) according to the manufacturer’s protocol. A control with no-RT was prepared in parallel, to confirm the absence of contaminating DNA in PCR experiments. An amount of 5 ng of initial RNA in RT reaction has been used to quantitatively evaluate the transcriptional levels of HERV-W *ENV*, HERV-K *ENV, N* SARS-CoV-2 [69] and *ACE2* genes by RT-qPCR (primer sequences are shown in Table 1A). The assays were performed in a StepOnePlus instrument (Applied Biosystems) using Platinum SYBR Green (Invitrogen, 11744-500). The housekeeping gene beta-2 microglobulin (*B2M*) was used to normalize the results in PBMC experiments whereas glyceraldehyde-3-phosphate dehydrogenase (*GAPDH*) was used in Vero cells experiments.

**Table 1:**
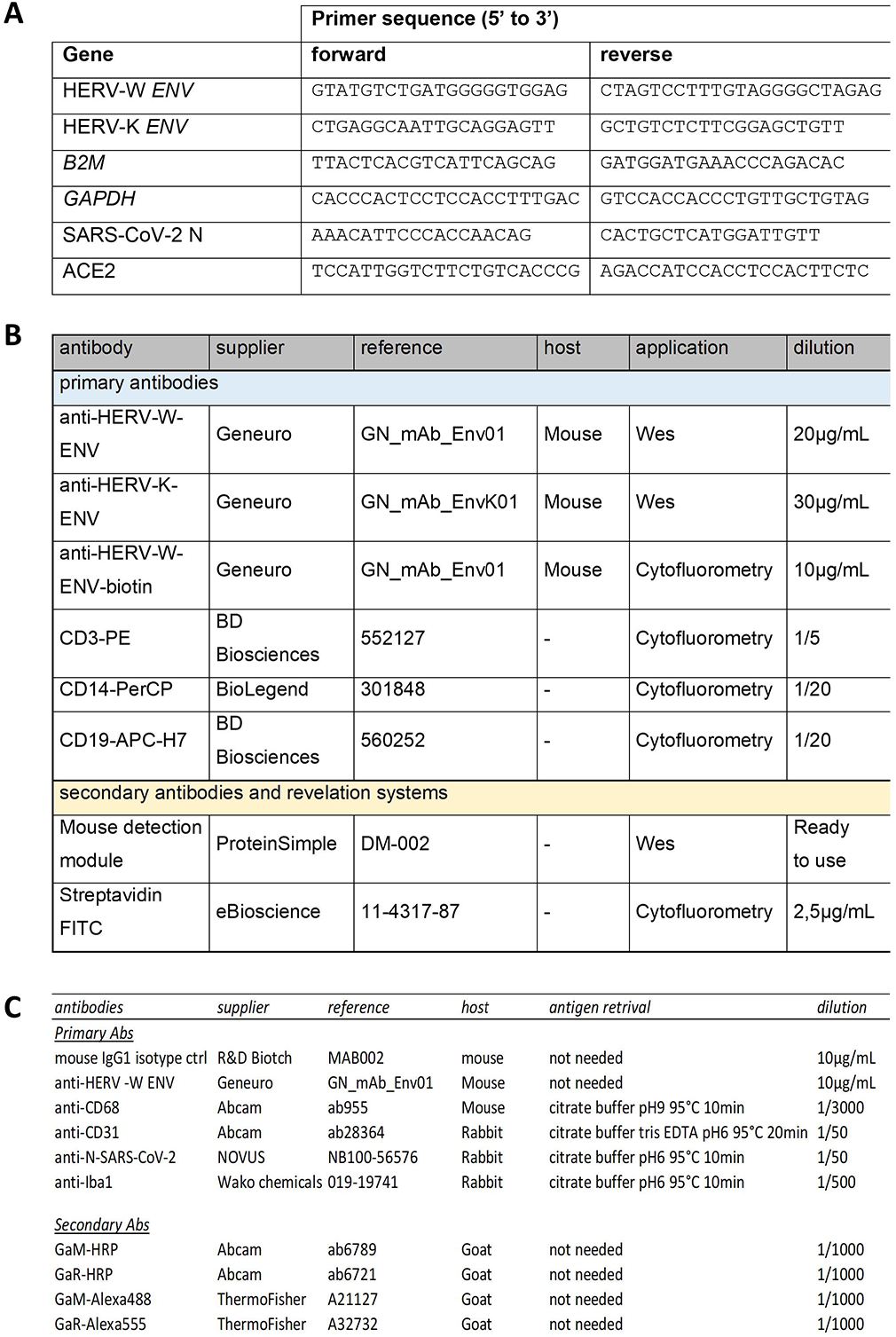
Primers sequences and antibodies. (A) Sequence of primers. (B) Antibodies used for Simple Western (Wes) and cytofluorometry. (C) Antibodies used for Immunohistochemistry and immunofluorescence.

Each experiment was completed with a melting curve analysis to confirm the specificity of amplification and the lack of any non-specific product and primer dimer in presence of the target. Quantification was performed using the threshold cycle (Ct) comparative method: the relative expression was calculated as follow: 2^-[ΔCt (sample) - ΔCt (calibrator)]^ = 2^-ΔΔCt^, where ΔCt (sample) = [Ct (target gene) – Ct (housekeeping gene)] and the ΔCt (calibrator) was the mean of ΔCt of (i) non infected/non treated cells for *in vitro* studies or (ii) PBMC of healthy controls for studies with patients.

#### A7. Quantification of Interleukin-6 (IL-6) secretion

IL-6 secretion was assessed in PBMC culture supernatant 2, 15 and 24 h after recombinant Spike exposure, by ELISA using BD Opt EIA Set Human IL-6 (BD, 555 220) according to supplier‘s recommendations.

#### A.8 Cell viability assay

PBMC viability was analyzed using CellTiter-Glo 2.0 Assay kit (Promega, G9241) according to the manufacturer’s protocol. Viability percentage was calculated using the following equation:

% of viability = (RLU_experimental_ – RLU_background_) / [Mean (RLU_control_ – RLU_background_)] x 100

Background: wells containing medium without cells; Control: untreated cells at 24 h post treatment.

### B. COVID-19 patients and healthy blood donors series

#### B1. PBMC, sera and plasma

Blood samples from healthy donors were obtained from the “Etablissement Français du Sang” (EFS) of Lyon (France). PMBC were isolated by Ficoll separation (Ficoll-Plaque PLUS) (GE Healthcare, 17-1440-02) and plasma were collected. Healthy donors signed a written Informed Consent Form, documented at the EFS, allowing the commercial use of their blood and blood components for medical research after definite anonymization.

PBMC and plasma of 27 (Lyon_2 cohort) and 20 (Lyon_3 cohort) SARS-CoV-2-positive individuals were obtained from the biobank of “Hospices Civils de Lyon” under conditions approved by the ethical committee (CRB, hôpital de la Croix-Rousse, Lyon France) (clinical data are provided in **Supplementary Tables S2 and S3**). Anonymized bioclinical and biological data were provided with samples. Upon receipt, PBMC were cultured 24 h in RPMI-1640 medium (Gibco, 61870-010) completed with 5% of decomplemented Human AB serum (Sigma, H4522).

Sera of 44 healthy controls (unknown COVID-19 status; prepandemic sampling), 43 SARS-CoV-2 PCR-negative patients with other diseases (OD) and 143 COVID-19 patients (SARS-CoV-2 PCR-positive) as well as anonymized clinical and biological data were provided as “Zaragoza cohort” by the “Biobanco del Sistema de Salud de Aragon” (PT20/00112), which in turn is integrated in the Spanish National Biobanks Network, Instituto de Salud Carlos III, Madrid, Spain (clinical data are provided in **Supplementary Table S4**). Samples and data from patients were processed following standard operating procedures. The provision of samples and the study protocol were approved by the scientific advisory board of the Biobank and by the local ethics committee (CEICA) (protocol C.P. - C.I. PI21/153 (07/04/21).

#### B2. Immunocapillary western-blot (Wes) detection of HERV-W ENV and HERV-K ENV in sera or plasma

For HERV-W ENV antigen, sera or plasma were incubated for 2 h at 25°C with gentle agitation in presence of 1X RIPA buffer (R0278-500ML, Sigma Aldrich) supplemented with 1% Fos cholin 16 (F316S-1GM, Anatrace) and protease inhibitor cocktail (5892791001, Roche). For HERV-K ENV antigen, sera or plasma were incubated 30 min on ice in extraction buffer containing 6M urea (GE Healthcare, 17-1319-01), 150 mM NaCl, 20 mM Tris pH 8, 0.6% IGEPAL® CA-630 (Sigma, I7771), 2mM EDTA, 1 mM PMSF and protease inhibitor cocktail (Roche, 0469313001). After 10 min of centrifugation at 10,000 x g, supernatants were collected. Deglycosylation was immediately performed and high molecular weight proteins were enriched on column filter as previously described [70].

HERV-W ENV and HERV-K ENV antigen detection was analyzed on the Wes device using Simple Western technology an automated capillary-based size sorting and immunolabeling system (ProteinSimple^TM^) as previously described [70]. The following primary antibodies were used: anti-HERV-W ENV mAb GN_mAb_ENV01 (-W01) (20 µg/mL) and anti-HERV-K ENV mAb GN_mAb_ENV-K01 (30 µg/mL) (antibodies description in Table 1B).

Signal quantification was made by calculating the complete area under the curve (AUC) defined by the curve (peak) above the baseline of the electropherogram, within the apparent molecular weight range of HERV-ENV deglycosylated antigen in this capillary matrix (350-450 KDa) using the Wes platform Compass^TM^ software (ProteinSimple, USA). Because this AUC also accounts for non-specific background signal within capillaries in this high molecular weight range, a cut-off of specificity for the quantification interval corresponding to the antigen detection is defined from the background signal generated by the same type of samples from negative healthy controls. The specificity threshold is statistically defined by the mean of negative controls (without possible outliers) + 2 standard deviations (cut-off= mean+2SD of negative controls). A relative quantification is therefore presented as a signal to noise ratio (S/N): Total peak AUC/cut-off =S/N. Positive antigen quantification corresponds to S/N values above 1, and negativity of the detection corresponds to values below 1. Since values below 1 do not represent a quantification of the specific signal due to the antigen, results with S/N<1 are all similarly negative and therefore normalized to the value S/N=1.

Samples were loaded in triplicates and the global signal containing the target hexamer HERV-W ENV (electrophoregram peak about 440 kDa) or HERV-K ENV protein (electrophoregram peak about 60-90 kDa) was measured. According to standard diagnostic rules for such immunoassays, in case of a single discordant value (CV>15%) was eliminated, keeping two consistent values (CV≤15%) of the triplicate for the calculation of the mean value as a result. In case of three discordant values with CV>15%, the result was deemed not interpretable.

#### B3. Cytofluorometry

PBMC isolated from COVID-19 patients and healthy donors were analyzed by cytofluorometry. 24 hours after PBMC seeding in culture medium as previously described, cells were pelleted by centrifugation. PBMC were incubated with FcR Blocking Reagent according to manufacturer’s protocol (Miltenyi Biotec, 130-113-199) and stained with CD3-PE (BD Biosciences, 552127), CD14-PerCP (BioLegend, 301848), CD19-APC-H7 (BD Biosciences, 560252) and 10 µg/mL GN_mAb_ENV01-biotin (GeNeuro, murine antibody). HERV-W ENV expression was revealed using Streptavidin FITC conjugate (eBioscience, 11-4317-87). Following cell surface staining, cells were fixed using Cytofix/Cytoperm kit (BD Biosciences, 554714) according to manufacturer’s instructions. Stained cells were acquired on a BD LSR Fortessa, fluorochrome emissions from the pool of antibodies were compensated using UltraComp eBeads plus Compensation beads (Invitrogen, 01-3333-42). Data were analyzed with FlowJo software (v.10). See table 1B for antibodies description.

### C. Immunohistology study

#### C1. Origin of tissue sections on slides

Paraffin embedded COVID-19 tissue slides (lung, heart, nasal and brain tissue) from 15 patient necropsies were provided by the National Institute of Respiratory Diseases in Mexico City, Mexico. The nasal/brain tissues sampling was performed via nasal cavity such as presented in Figure 8A (Ethical committee approval and legal authorization: Autorización para realizar estudios postmortem INER-SAM-01, Secretaria de Salud, Mexico). Paraffin embedded tissue sections of non-COVID-19 lung (normal appearing tissue from lung cancer) from 3 patients were ordered at the biobank from the biological research center (CRB) of Croix-Rousse hospital, Lyon, France. Samples description and clinical data are respectively provided in **Supplementary Tables S5 and S6.**

#### C2. Immunochemistry (IHC)

Paraffin tissue sections were rehydrated by successive bath of toluene, degreasing concentration of ethanol and 1X PBS. Depending of the antibody/antigen couple, an antigen retrieval step can be needed consisting to boil samples in citrate buffer at appropriate pH (condition detailed in the Table 1C). Endogenous peroxydases were inhibited in a 30 min bath of 4% H_2_O_2_. Permeabilization was performed during 5-10 min in 1X PBS+0.2% Tween 20 and the non-specific sites interaction were blocked by an incubation in 3% horse serum in 1X PBS+0.2% Tween 20 during 30min at RT. Details of primary and secondary antibodies (all diluted in blocking buffer) concentrations were detailed in Table 1C. Primary antibodies were incubated overnight at 4°C and secondary antibodies were incubated during 45 min at RT. Revelation was performed using AEC kit (Vector, SK-4200) following supplier indications. Nuclei were counter-stained during 3 min with Harris hematoxylin (filtrated and 3-fold diluted) before a quick rinse in water. Slides were mounted using Fluoromount (Southern Biotech). IHC and IF slide observation and image acquisition were performed on NIKON Eclipse TS2R microscope and analyzed using ImageJ software.

## Results

### 1. Exposure to SARS-CoV-2 virus triggers HERV-W and -K *ENV* mRNA early transcription in PBMC of heathy donors

We initially analyzed whether infectious SARS-CoV-2 could modulate the expression of HERV-W and HERV-K *ENV* genes in lymphocytes from healthy blood donors (HBD). PBMC of blood donors were cultured with or without infectious SARS-CoV-2 and RNA was collected at 2h post-inoculation. In PBMC from 3 out of 11 donors (27%), HERV-W *ENV* RNA levels were increased after exposure to wild type SARS-CoV-2 virus (Figure 1A). The same donors also showed relative transcriptional activation of HERV-K *ENV* and a low response for HERV-W only was also seen in another donor (D27).

**Figure 1:**
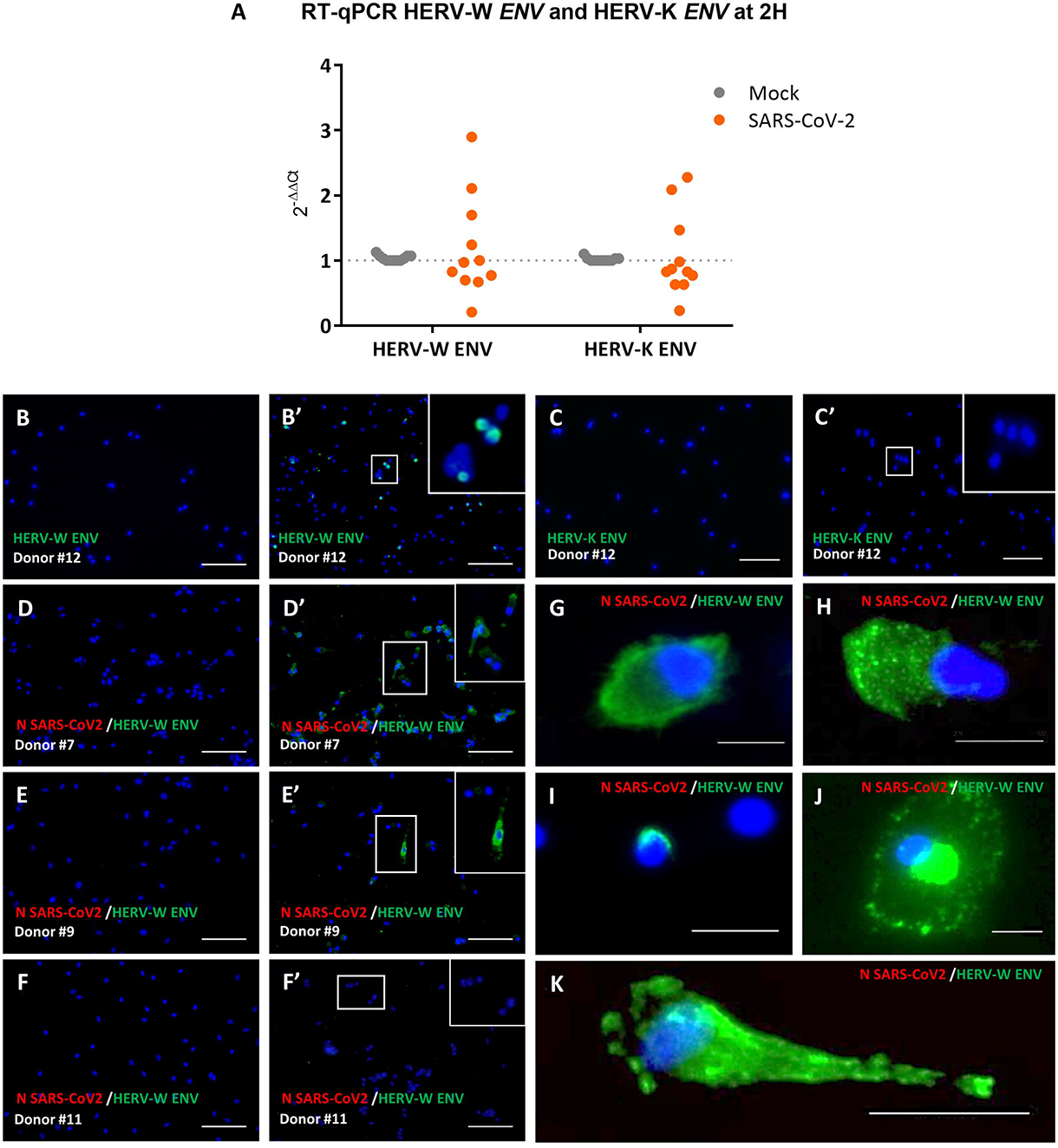
HERV-W *ENV* and HERV-K *ENV* RNA quantification and HERV-W ENV protein immunodetection in PBMC from healthy blood donors exposed to SARS-CoV-2. (A) The level of mRNA of HERV-W *ENV* and HERV-K *ENV* of PBMCs cultures from 11 healthy blood donors, exposed or not to infectious SARS-CoV-2 (MOI:0.1), was analyzed by RT-qPCR. The graph presents mean results from monoplicate (4 out 11 donors) and triplicate (7 out 11 donors) cultures at 2h post-exposure. Numbers of donors with increased expression of HERV-W and HERV-K are indicated and values of each sample is presented on the **Supplementary table S1**. (B-K) PBMC cultures of 8 healthy blood donors, inoculated or not with SARS-CoV-2 at 0.1 MOI, were collected at either 3 or 7 days post-exposure For each culture, GN_mAb_Env01 and anti-N-SARS-CoV-2 antibodies were used respectively to detect HERV-W ENV (green labelling) and nucleocapsid protein of SARS-CoV-2 (red labeling). (B-B’ and C-C’) PBMCs from healthy blood donor #12 were inoculated (B’-C’) or not (B, C) with SARS-CoV-2 virus (MOI: 0.1). GN_mAb_Env01 (B, B’) or GN_mAb_Env-K01 (C, C’) antibodies were respectively used to detect HERV-W ENV or HERV-K ENV (green labelling). (D-F’) PBMC cultures from 3 representative donors with variable number of positive cells: Donor # 7 at day 7 (D, D’) and Donor # 9 at day 3 (E, E’) and a non-responding culture from Donor # 11 at day 7 (F, F’). The different morphological aspects of HERV-W ENV positive cells are presented with high magnifications (G-K). DAPI was used to stain nuclei (blue staining). Bars B-F’ = 100µm; Bars G-K = 25µm.

Further kinetics analysis showed that in PBMC of 4 representative donors, the level of both HERV-W *ENV* and HERV-K *ENV* RNA was decreased at 19h and 24h post-inoculation to a level even below the one detected in sham-inoculated control cultures from the same donors at the same time points (**Supplementary Figure S1 A-B**).

Interestingly, a similar decrease of HERV-W and -K *ENV* RNA levels was observed from the earliest time point in PBMC from the “non-responding” donors, i.e., after 2h exposure to SARS-CoV-2 virus.

Analysis of SARS-CoV-2 *N* RNA kinetics only showed abundant RNA load from the inoculum with a significant decrease at 19h post-infection (inoculation), which confirmed the absence of viral replication in PBMC (**Supplementary Figure S1 C**).

### 2. Exposure to SARS-CoV-2 triggers HERV-W envelope (ENV) production from PBMC of heathy donors

We next analyzed whether the HERV RNA expression was followed by HERV protein production. PBMC cultures of independent blood donors, inoculated with SARS-CoV-2 or sham-inoculated, were collected at either 3 or 7 days post-exposure. The presence of envelope proteins of HERV-W, HERV-K or SARS-CoV-2 N protein was analyzed by immunofluorescence (Figure 1 B-K). HERV-K ENV staining was not detected in cells after exposure to SARS-CoV-2, even when HERV-W ENV protein was detected in the same cells (Figure 1 B-C’). HERV-W ENV expression was donor-dependent as observed in 3 out of 8 HBD PBMC cultures inoculated with SARS-CoV-2. It was abundantly detected in donor #7 (Figure 1D’), moderately in #12 (Figure 1B’), and in isolated cells of #9 (Figure 1E’), whereas not detected in others (Figure 1F**’,** donor #11), similarly to “non-responders” with previous mRNA profiles. Nonetheless, in HERV-W ENV positive cells of “responders”, a marked expression was seen at high magnification (Figure 1 G-K). SARS-CoV-2 antigen was not detected in any of the conditions. In all immunofluorescence analyses, neither HERV-W ENV, nor HERV-K ENV protein expression was detected in control cultures without SARS-CoV-2 (Figure 1 B, C, D, E **and** F).

Parallel immunofluorescence analysis showed clear expression of SARS-CoV-2 N and Spike proteins in Vero-E6 cells used as a positive control (**Supplementary Figure S2 A-D**). The specificity of the anti HERV-W antibodies was also confirmed with the absence of staining in the same SARS-CoV-2-infected Vero-E6 cells, with increased SARS-CoV-2 *N* mRNA and negative RT-qPCR for HERV-W *ENV* and HERV-K *ENV* (**Supplementary Figure S2 E-G**). Positive controls for anti-HERV antibodies sensitivity and specificity on cells transfected with expression plasmids for HERV-W (MSRV strain versus Syncytin-1) or -K ENV antigens (**Supplementary Figure S2 H-N**) and negative control staining with secondary antibodies (**Supplementary Figure S2 O-U)** were performed in parallel. The specificity of the anti-HERV-W ENV antibody was already established with different conditions in a previous study [70].

### 3. Exposure to SARS-CoV-2 triggers HERV-W envelope production in T-cell subsets of heathy individuals

HERV-W ENV protein expression having been observed in CD3^+^ T-cells of COVID-19 patients [60], we analyzed whether direct exposure to SARS-CoV-2 could induce HERV-W ENV in T lymphocytes from healthy donors. We analyzed HERV-W ENV expression in PBMC cultures from three healthy donors, with or without exposure to SARS-CoV-2, using cytofluorometry analysis on non-permeabilized cells. As illustrated in Figure 2, CD3^+^ T lymphocytes were identified with the gating strategy in non-infected (NI) cultures. CD3+ cells comprised CD3^high^ T-cells, physiologically representing naïve/non-activated cells, and CD3^+^ cells with increased size, normally representing activated T-cells following, e.g., antigenic stimulation, further decreasing CD3 exposure at heir surface and corresponding to the CD3^low^ subpopulation [71, 72]. Furthermore, in order to avoid confusion between CD14+ monocytes and larger CD3^low^ T-cells, we have evaluated T-cell populations by eliminating CD14+ cells and gating specifically CD14-cells (Figure 2A-D). Indeed, when inoculated with SARS-CoV-2, CD3+ cells showed HERV-W ENV cell-surface expression. Minimal fluorescence associated with HERV-W ENV detection (compatible with technical noise) was observed in sham-inoculated CD3^+^ T-cells (Figure 2A), whereas a significant increase was characterized in SARS-CoV-2-associated cultures mainly observed in CD3^low^ cells (Figure 2B). In the presence of SARS-CoV-2, CD3^low^ cells may also reflect the predicted superantigenic properties of the Spike protein [73]. Double-labeling with anti-CD3 and anti-HERV-W ENV specific antibodies indicated that a significant proportion of CD3^low^ T-cells was positive for HERV-W ENV at the cell-surface both at 24h and at 72h after exposure to the virus (Figure 2A and 2C, CD3^+^ top panels) compared to CD3^high^ T-cells (Figure 2B and 2D, CD3^+^ bottom panels). Sham-infected cultures did not express notable levels of HERV-W envelope antigen in CD3^+^ T-lymphocytes (Figure 2A and 2C, CD3^+^ top and bottom panels). The percentage of HERV-W ENV positive cells in each condition is represented in Figure 2E and 2F at 24h and 72h post-inoculation with SARS-CoV-2.

**Figure 2:**
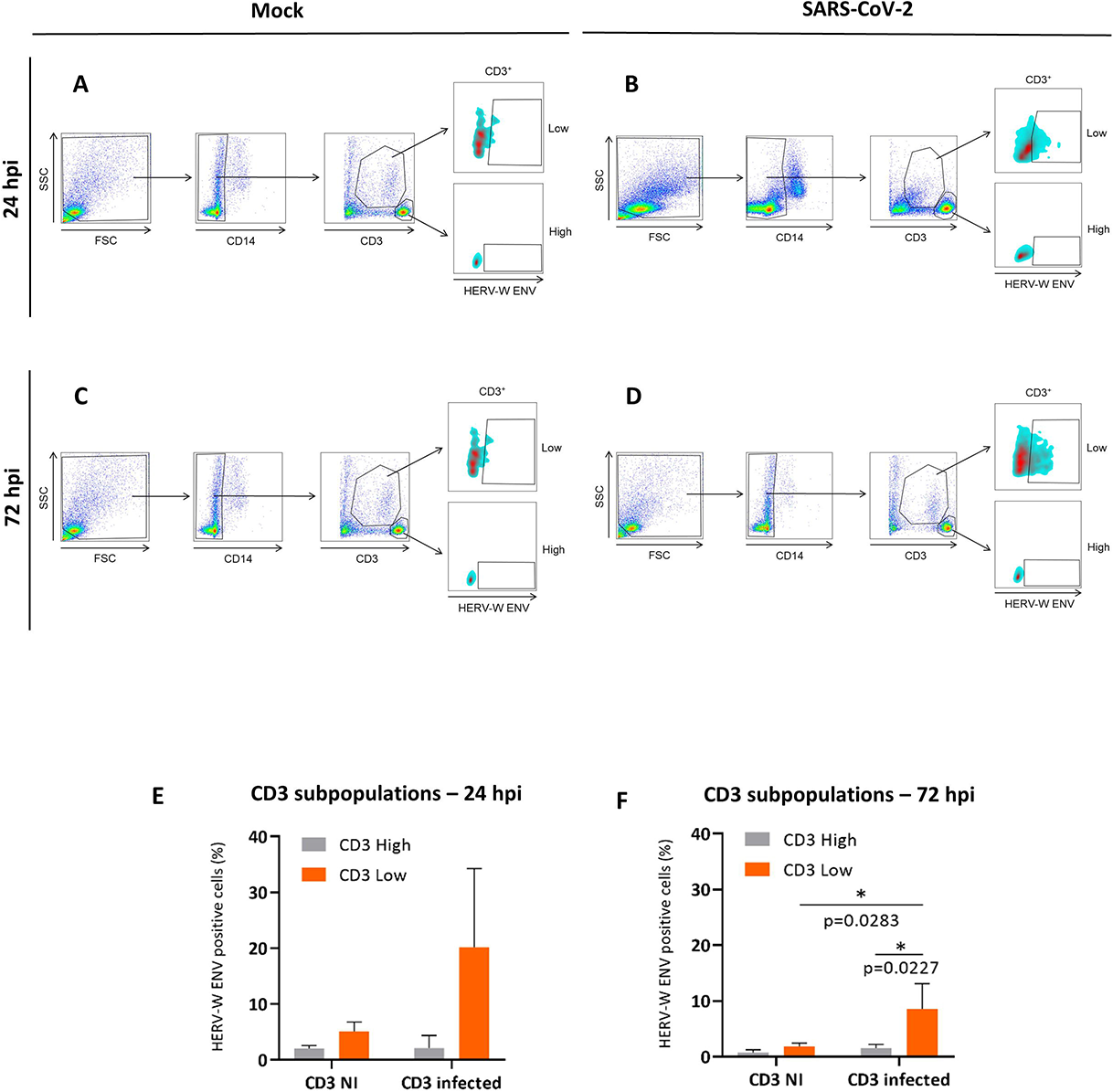
SARS-CoV-2 induces the expression of HERV-W *ENV* in CD3^low^ T cells. PBMCs from 3 healthy donors were either incubated with SARS-CoV-2 (MOI=0.1) for 24h (B) or 72h (D), or remained unexposed to the virus (A, C). Cells were stained using anti-CD3, anti-CD14 or GN_mAb_Env01 antibodies and analyzed by flow cytometry. The percentage of HERV-W ENV positive cells in CD14 and CD3 T cells, with identification of CD3^high^ and CD3^low^ T cell subpopulations (after suppression of CD14 cells from the gating) was determined (A-D). The percentages of HERV-W ENV positive cells from subpopulations within CD3 T lymphocytes at 24h post-infection and at 72h post-infection (pi; average from 3 donors ±SD) are presented with histograms (E, F). Statistical analysis was performed using Tukey’s multiple comparisons test. *: p<0.05.

### 4. Exposure to SARS-CoV-2 recombinant trimeric Spike protein triggers HERV-W ENV protein production in PBMC of some heathy individuals

We previously observed a rapid response to SARS-CoV-2 virus characterized by an early peak of RNA followed by HERV-W ENV protein expression in PBMC maintained in culture (≤5 days; Figures 1-2). This immediate RNA response in the absence of detectable infection by SARS-CoV-2, prompted us to investigate a possible direct stimulation by SARS-CoV-2 proteins, particularly by its surface Spike protein.

A recombinant trimeric Spike protein without stabilizing mutations [74], was added into the culture medium of PBMC (Figure 3).

**Figure 3:**
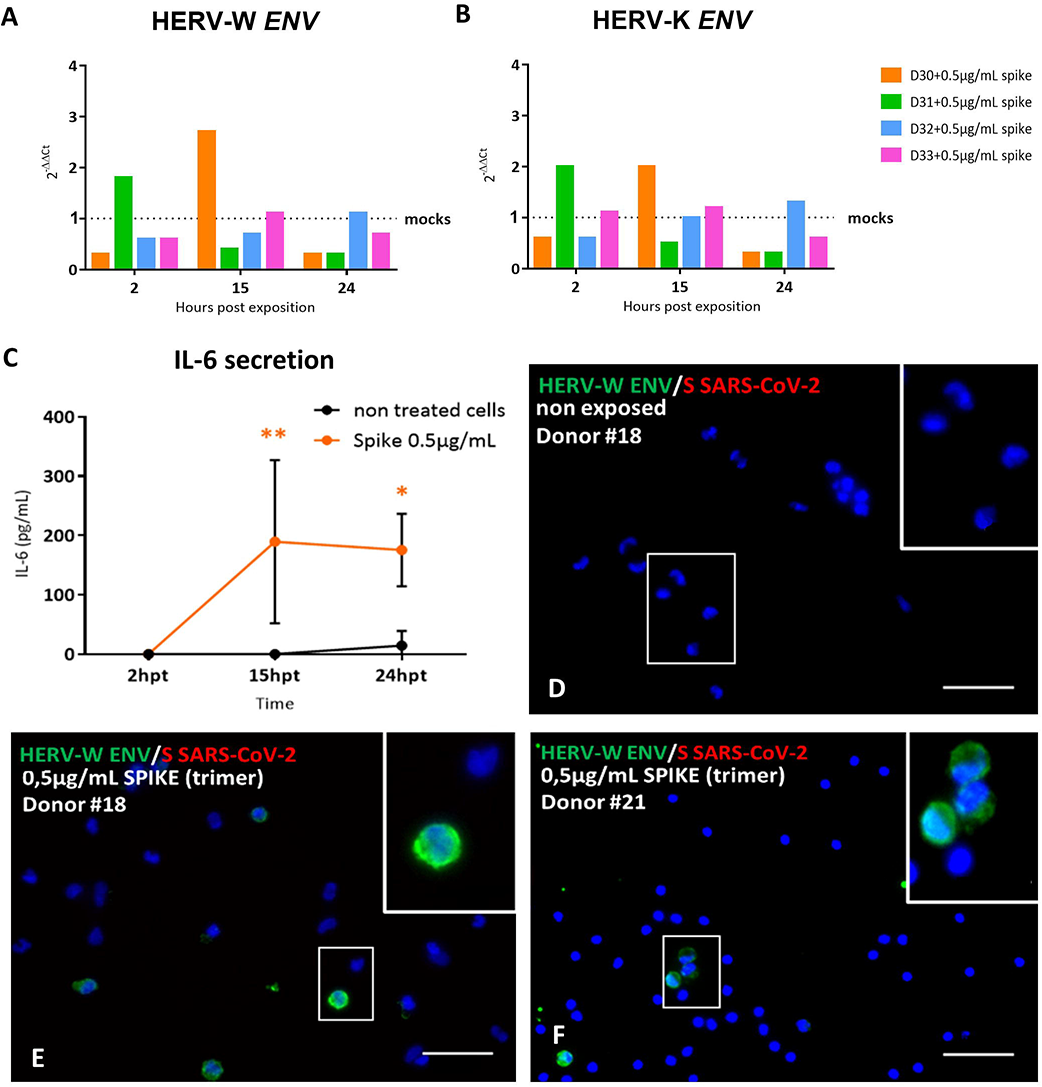
HERV-W *ENV* and HERV-K *ENV* mRNA induction after exposure to SARS-CoV-2 Spike trimer, followed by IL-6 secretion and HERV-W ENV protein expression in PBMC from healthy donors. (A-B) PBMCs from 4 healthy blood donors were exposed to 0.5 µg/mL of active trimer Spike recombinant protein in parallel to mock-control (buffer) and HERV-W *ENV* (A) and HERV-K *ENV* (B) mRNA levels were assessed at 2h, 15h and 24h post-inoculation (pi) by RT-qPCR using specific primers. Results were presented as the fold change of the corresponding non-infected condition. (C) PBMCs isolated from 3 healthy blood donors (donors #30 to 32) were incubated, or not, with 0.5 µg/mL of recombinant spike trimer. IL-6 secretion was monitored in culture at 2, 15 and 24h post treatment (hpt) using the BD® Opt EIA Set Human IL-6® ELISA dosage. Statistical analysis: Sidak’s multiple comparison. Here, mock-treated and treated condition were compared at each post-treatment time-point 2 h, 15 h and 24 h (*: 0.5<*p*<0.01; **: 0.01<*p*<0.005). (D-F) PBMCs from healthy blood donors were exposed during 24 h with 0.5 µg/mL of active trimer Spike recombinant protein. Here, results obtained on 2 responding donors (donor # 18 (E) and donor # 21 (F)), are presented. (D) Untreated PBMCs from the responding donor # 18 are also shown as non-exposed mock control. They did not present any HERV-W ENV detection using GN_mAb_Env01 antibody (green staining) and no signal corresponding to the recombinant Spike protein was detected with anti-S-SARS-CoV-2 antibody (red staining). (E-F) HERV-W ENV was detected in few cells of cultures exposed to the spike trimer (green staining). Higher magnifications of HERV-W ENV positive cells are presented in white squares. DAPI was used to stain nuclei (blue staining). Bars D-F = 50µm.

PBMC RNA was collected at 2h, 15h and 24h post-inoculation and analyzed by RT-qPCR for HERV-W and HERV-K envelope gene expression. Donor #31 showed increased RNA levels for both HERV-W and HERV-K *ENV* at 2h post-inoculation. Donor #30 showed a peak of HERV-W and HERV-K *ENV* RNA at 19h, while both RNA levels remained rather stable or decreased in donors #32 and #33 (Figure 3 A-B). RNA levels for both HERV-W and HERV-K *ENV* also decreased below the baseline of identical non-exposed cells after having shown a peak of transcription, a pattern similar to the one observed in Figure 1. Of interest, donor #30 was the only one tested positive for anti-SARS CoV-2 serum antibodies (data not shown), which did not prevent HERV transcriptional activation by this recombinant spike trimer but coincided with delayed peak of HERV RNA.

IL-6 secretion was significantly increased after exposure to SARS-CoV-2 S antigen in PBMC from all tested donors, including those not responding in terms of HERV activation as illustrated with the ones presented in Figure 3C. Thus, HERV activation occurred independently from IL-6 production and, beyond RNA analysis, HERV-W ENV protein production was confirmed by immunofluorescence analysis at 72h in cultured PBMC of 2 out of four other donors inoculated with spike trimers (0.5 µg/mL) and not in mock-control cultures (Figure 3 D-F). HERV-W ENV positive cells were detected, as previously seen with the infectious virus. Moreover, a relative increase in HERV-W ENV RNA was quantified, despite a low proportion of activated cells within the cultures. Of interest, the absence of ACE2 expression in PBMC (**Supplementary Figure S3 A**) suggests an interaction of the spike protein with another receptor.

Cell viability was measured and did not vary significantly within the culture period of experiments (≤5days; **Supplementary Figure S3 B**).

### 5. HERV-W ENV protein is expressed at the surface of T-lymphocytes from COVID-19 patients and correlates with the detection of soluble HERV-W ENV hexameric antigen in plasma

After our initial data has demonstrated that that the exposure to SARS-CoV-2 induces the expression of HERV-W in PBMCs of certain healthy donors, we further analyzed the levels of HERV-W ENV antigen expressed on PBMC and compared it with the detection of soluble antigen in plasma from a cohort of hospitalized COVID-19-positive patients. Biological and clinical data of these patients from Hôpital de la Croix-Rousse, Lyon, France are presented in **Supplementary Table S2**.

The proportion of non-permeabilized HERV-W ENV positive cells, i.e., with antigen at the cell surface, from 27 patients with COVID-19 compared to 14 HBD is presented in Figure 4. Examples illustrating the gating conditions by cytofluorometry are presented in **Supplementary Figure S4**. Similar gating strategy aiming at eliminating potential confusion between monocytes and larger T-cells was applied by gating CD14-cells to specifically study CD3+ T lymphocytes, and CD14+ to specifically investigate monocytes (**Supplementary Figure S4A and C**). As shown, a highly significant difference was observed between COVID-19 patients and HBD controls on CD3^+^ T-cells (p=0.0005 for CD3^low^ T-cells and p=0.0023 for CD3^high^ T-cells) (Figure 4A and 4B). However, while the amount of CD3^high^ T-cells positive for HERV-W ENV was low but significant when compared to HBD (p=0.0005) (Figure 4A), significant but much higher levels were seen in CD3^high^ T-cells of COVID-19 individuals (p=0.0023 Vs HBD; Figure 4B). Interestingly, whereas the levels of CD14^+^ monocytes stained for HERV-W ENV remained equivalent between HBD and COVID-19 patients (Figure 4C), a significant difference was also observed in CD19^+^ B-cells between HBD and COVID-19 groups (p=0.031) (Figure 4D).

**Figure 4.**
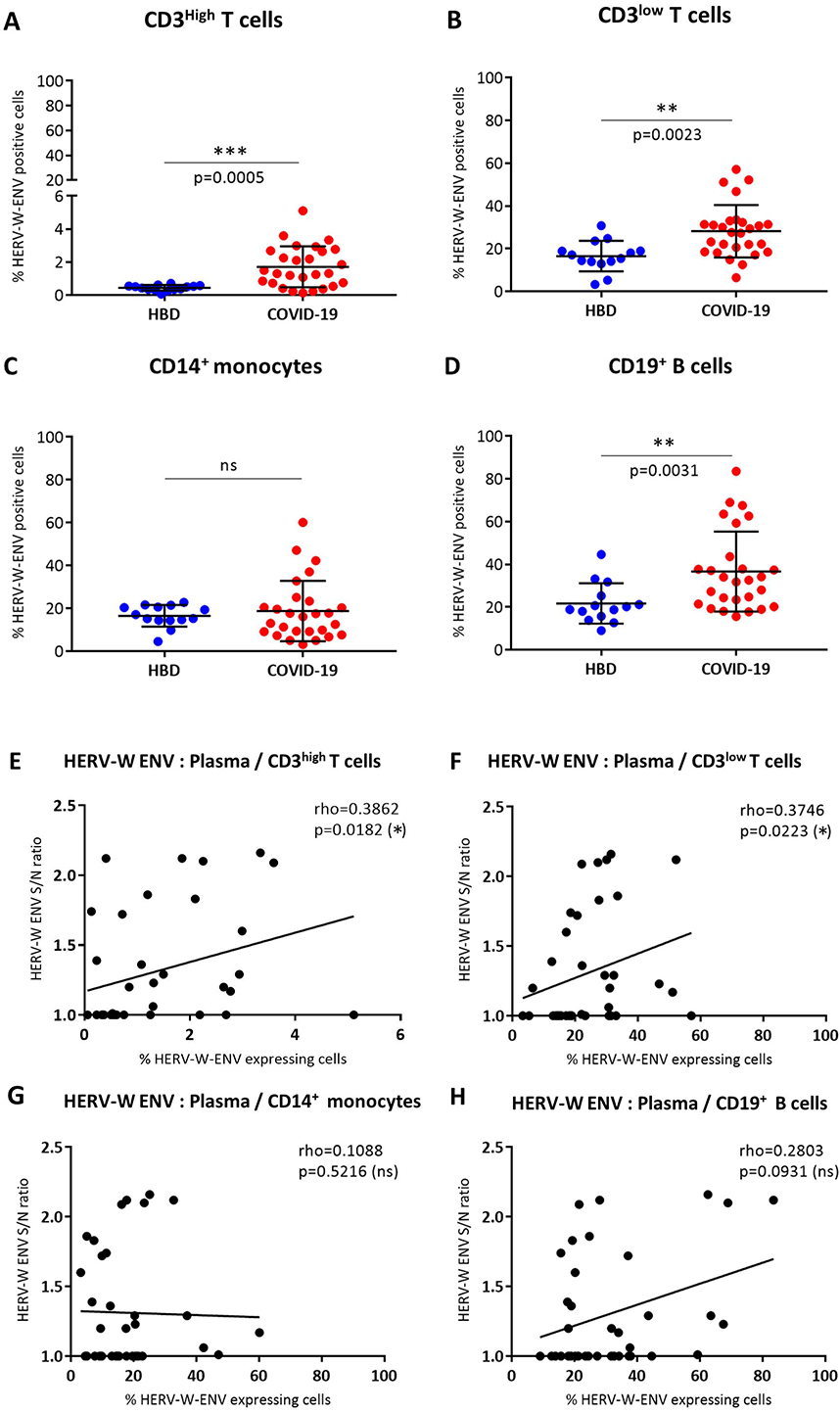
Production of HERV-W ENV in PBMC of COVID-19 patients and correlation with the plasma concentration of soluble HERV-W ENV. (A-D) The percentage of HERV-W ENV positive cells was analyzed by cytofluorometry in PBMC from 14 HBD (blue dots) and 27 COVID-19 patients (red dots). The percentage of HERV-W ENV positive cells has been evaluated in CD3^high^ T cells (A), CD3^low^ T cells (B), CD14^+^ monocytes (C) and CD19^+^ B cells (D). Unpaired t test (A, B) and non-parametric Mann-Whitney test (C, D) were used to compare HBD and COVID-19 groups (ns: *p*>0.5; *: 0.5<*p*<0.01; **: 0.01<*p*<0.005). (E-H) Scatter plots presenting the quantification of the soluble HERV-W ENV in plasma (Y axis) and the percentage of HERV-W ENV positive CD3^high^ (E), CD3^low^ (F), CD14+ monocytes (G) and CD19^+^ B cells (X axis) in blood of 14 HBD and 27 COVID-19 patients. Gating strategy for cytofluorometry analyses is presented in **Supplementary Figure S4.**

HERV-W ENV soluble hexameric antigen was also detected in plasma of COVID-19 patients and its correlation with HERV-W ENV expression levels previously determined by cytofluorometry on each cell population was assessed (Figure 4 E-H). A moderate but significant correlation was observed between the HERV-W ENV antigen released in plasma and the one expressed on the membrane of both naïve CD3^+^ and activated CD3^+^ T-lymphocytes (p<0.02 and p<0.03, respectively) (Figure 4E and 4F). In parallel, no correlation was found for CD14^+^ monocytes (Figure 4G) and CD19^+^ B-cells (Figure 4H) between cytofluorometric and plasma analyses. In addition to the previously reported correlation of HERV-W ENV expression quantified by RT-qPCR with the expression determined by cytofluorometry in CD3^+^ T-lymphocytes from COVID-19 patients [60], the present correlation (Figure 4F) provides another orthogonal validation of HERV-W ENV antigen detection by two unrelated techniques in COVID-19 blood samples.

Finally, SARS-CoV-2 IgG serology for multiple antigens was also analyzed, but no significant correlation was observed between anti-SARS-CoV-2 IgG titers and HERV-W ENV hexamer antigenemia in plasma samples (**Supplementary Figure S5 A**).

### 6. Detection of soluble HERV-W ENV antigen in plasma from hospitalized COVID-19 patients: correlation with biological and clinical parameters

HERV-W ENV hexameric antigen was further quantified by immunocapillary analysis (digital Wes platform) in plasma of a second series of patients presenting with different severity of COVID-19 symptomatic forms, also caused by different SARS-CoV-2 variants in patients who were also affected by various pre-existing diseases (Hopital de la Croix Rousse, Lyon-France; Cf. **Supplementary Table S3**).

As shown in Figure 5 A-C, HERV-W ENV soluble antigen was significantly detected in plasma from COVID-19 patients (11/21) compared to healthy blood donors (0/11). HERV-K ENV antigen was not detected in any plasma sample (**Supplementary Figure S5 B-C**). Patients were subsequently classified with the clinical scale for COVID-19 recommended by the National Institute of Health of the USA guidelines (https://www.covid19treatmentguidelines.nih.gov/overview/clinical-spectrum/). As presented in Figure 5D, the mean titers of HERV-W ENV progressively increased with disease severity and a positive detection was observed in all severe forms. The comparison between the ‘severe’ group and that of healthy controls was statistically significant despite low numbers (Mann Whitney U-test, p=0.0037).

**Figure 5.**
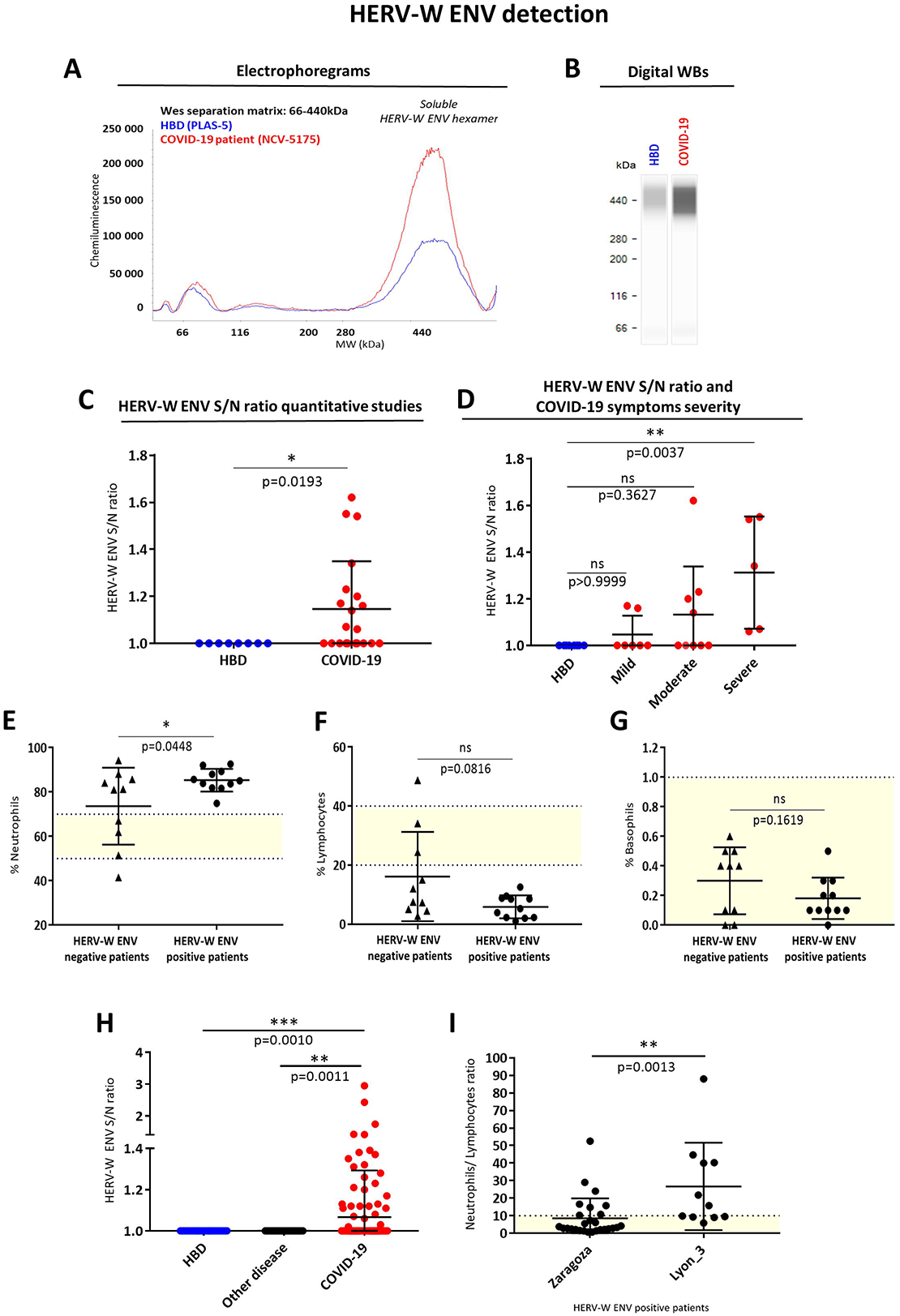
Detection of soluble HERV-W ENV in plasma from COVID-19 patients and correlation with blood biomarkers. (A-D) Immunodetection of HERV-W ENV in plasma samples from 11 HBD and 21 COVID-19 patients from Lyon (France, “Lyon_3 cohort”) after protein extraction and Wes analysis under denaturing conditions with primary antibody GN_mAb_ENV01. Electrophoregrams (A) and digital western blots (B) representations of representative HBD (blue panel) and COVID-19 patient (red panel). (C-D) AUC of the HERV-W ENV hexamer (electrophoregram peak about 440 kDa) was measured and expressed as HERV-W ENV S/N ratios. S/N ratio below the positivity threshold (< 1) were corrected to a value equal to 1 (negative detection). (C) HERV-W ENV S/N ratios of HBD (blue dots) and all COVID-19 patients (red dots). Statistical significance was determined by non-parametric Mann-Whitney test (*: 0.5<*p*<0.01). (D) HERV-W ENV S/N ratios of HBD (blue dots) and COVID-19 patients with mild (orange dots), moderate (red dots) or severe (dark red dots) symptoms. Significance determined by Dunn’s multiple comparisons test. (E-G) Scatter plots comparing 21 COVID-19 patients either with positive HERV-W ENV antigen detection in plasma (X axis; S/N>1) (and or with negative detection (X axis; S/N ≤1,normalized to 1) for their corresponding blood percentage in blood cell counts of Neutrophil Polymorphonuclear cells (E), of Lymphocytes (F) and Basophil Polymorphonuclear cells (G) (Y axis). Statistical significance was determined by Unpaired t test (E, G) and non-parametric Mann-Whitney test (F) (n.s: *p*>0.5; *: 0.5<*p*<0.01). Yellow areas represent the reference range for blood tests in healthy adults (QC-validated normal values provided with results). (H) Detection of soluble HERV-W ENV in serum from COVID-19 patients at the day of RT-qPCR diagnosis (Spain, “Zaragoza cohort”). HERV-W antigenemia, related to HERV-W ENV S/N ratios in plasma or sera, of HBD (blue dots, n = 44), of non-COVID-19patients (“other disease”, black dots, n = 43) and of COVID-19 patients (red dots, n = 143) are compared using statistical Dunn’s multiple comparison test (**: 0.01<*p*<0.005; ***: 0.005<*p*<0.0001). (I) Neutrophils/lymphocytes ratio comparison between patients with HERV-W ENV positive antigenemia (S/N>1) from early and late COVID-19 cohorts consisting in: (i) outpatients from the last cohort with early post-PCR COVID-19 diagnosis cases from Zaragoza biobank, Spain and, (ii) inpatients from the last cohort of hospitalized COVID-19 patients from Lyon, France. The normal interval for the neutrophils/lymphocytes ratio is represented by the colored area above the X axis. Statistical significance was determined by Mann-Whitney U-test (**: 0.01<*p*<0.005). HBD: Healthy Blood Donor. S/N : Signal/Noise ratio.

Analysis of the biological data also revealed a significant difference (Mann Withney U-test, p=0.0448) between polymorphonuclear neutrophils counts of patients with positive HERV-W ENV antigenemia (S/N>1) and of those without (S/N<1; Figure 5E). Interestingly, all neutrophils counts from patients with positive antigenemia (11/11) were above the physiological interval (Figure 5E). In parallel, they all had lymphopenia (Figure 5F), though not statistically different from other COVID-19 cases without detectable HERV-W antigenemia. No particular difference was observed for other white blood cells, as shown for example with basophils (Figure 5G).

### 7. Detection of soluble HERV-W ENV antigen in serum from outpatients at first diagnosis with SARS-CoV-2 PCR detection

To study HERV-W ENV antigenemia in patients representing a wider range of clinical status at early COVID-19 diagnosis (compared to patients with other diseases and healthy controls-HC) its soluble antigen was quantified in sera from a third cohort consisting in two successive sampling series of SARS-CoV-2 PCR positive cases with heterogeneous clinical presentation (described in **Supplementary Table S4**). As shown in Figure 5H, a significant difference was found between all COVID-19 patients and HBD or other diseases (Mann Whitney U-test, respectively p=0.0010 and p=0.0011). 21% of COVID-19 sera were positive for HERV-W ENV among the 144 SARS-CoV-2 PCR positive cases, while none of the 44 PCR negative HBD were positive for HERV-W ENV (Chi^2^=8.796, p<0.001). Similarly, HERV-W ENV antigen was not detected in 43 sera from non-COVID-19 other diseases either (Chi^2^=8.604, p<0.001).

Finally, the ratio between neutrophils and lymphocytes counts (N/L), previously suggested as a biomarker of COVID-19 severity [23], was significantly increased in COVID-19 cases with HERV-W ENV positive antigenemia. This is illustrated in Figure 5I with samples from the previous cohort of hospitalized patients (“Lyon-3” series, Table S3 and Figures 5C and 5D) with more symptomatic and longer disease evolution, when compared with the cohort of early diagnosed outpatients (series from Zaragoza, **Table S4 and** Figures 5H). These results also highlight the difference in the clinical status of patients between the two cohorts and, possibly, the relevance of an early HERV-W ENV positive antigenemia.

### 8. Expression of SARS-CoV-2 and HERV-W ENV antigens in tissues from autopsies of acute COVID-19 patients

To better understand the consistency and magnitude of HERV-W ENV expression in SARS-CoV-2-infected patients, we next analyzed the expression of this antigen in tissues from COVID-19 patients. Post-Mortem lung, heart, nasal mucosa and brain tissue samples were obtained from patients deceased from severe acute forms of COVID-19 (**Supplementary Table S5**).

Lung tissue staining with monoclonal antibodies targeting SARS-CoV-2 Nucleocapsid (N-Protein) or HERV-W ENV is presented in Figure 6. SARS-CoV-2 antigen was readily detected in lung epithelial cells but not in alveolar macrophages (Figure 6A). Conversely, HERV-W ENV antigen was strongly expressed at the cell membrane of macrophages but not in neighboring SARS-CoV-2 positive pneumocytes (Figure 6B). HERV-W ENV positive cells were confirmed to be alveolar macrophages with CD68 staining in an adjacent slide (Figure 6C). The specificity of both SARS-CoV-2 N and HERV-W ENV proteins was confirmed by the absence of staining within similar sections of lung tissue from non-COVID controls, i.e., normal tissue surrounding lung tumors (Figure 6 A’-B’ **and Supplementary Figure S6)**. Wider areas of COVID-19 lung tissue stained with for HERV-W ENV are presented in Figure 6 D-F, which show HERV-W ENV expression in scattered infiltrating lymphoid cells (Figure 6F and 6 G-I) and sub-epithelial strong staining of cellular aggregates (Figure 6I and 6J). The specificity of the detection antibody was further assessed in the absence of staining with an isotype control in the same tissue samples from COVID-19 (Figure 6 **D’-F’)**. HERV-W ENV positive cells are presented with higher magnification in Figure 6 G-J, which corresponds to massive infiltrates of lymphoid cells (Figure 6G) also diffusing within alveola (Figure 6H), and to aggregated cells or clots within blood vessels (Figure 6 I-J). Positive endothelial cells were also seen (Figure 6I) and an isolated blood clot with strong HERV-W ENV staining (Figure 6J) represents a circulating micro-thrombus.

**Figure 6:**
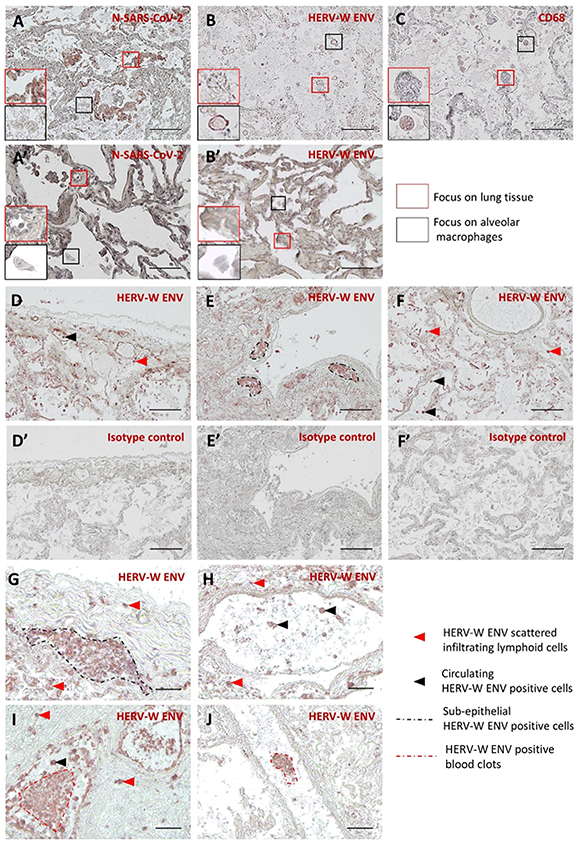
Immunohistological detection of SARS-CoV-2 and HERV-W ENV antigen in post-mortem lung tissue from acute COVID-19 cases. N-SARS-CoV-2 (A, A’) and HERV-W ENV (B, B’) immunodetection (brown-red staining) were performed on lung tissue sections from necropsies of COVID-19 patients (A-C) and non-COVID-19 (A’, B’, normal appearing tissue from lung cancers). In order to identify the HERV-W ENV producing cell type, a CD68 immunostaining, targeting a specific marker of macrophages, was performed on lung sections from COVID-19 lungs (C). Higher magnifications of pneumocytes (A-C, red squares) and alveolar macrophages (A-C, black squares) are presented on COVID-19 and non-COVID-19 tissues for comparison at the same magnification. HERV-W ENV immunodetection specificity was also assessed on COVID-19 tissues by comparing anti-HERV-W ENV murine IgG1 GN_mAb_Env01 (D-F) and murine IgG1 isotype control (D’-F’) at the same concentration (10 µg/mL) on adjacent tissue sections. Specific HERV-W ENV staining on COVID-19 lung showed several HERV-W ENV positive cell types, not seen in all slides, such as small round-shaped infiltrated cells with lymphoid morphology (G), isolated cells within large blood vessel lumen (H), blood vessel endothelium cells (I) or very small irregular cells aggregated in blood vessels compatible with platelet morphology (J). Red arrowhead: HERV-W ENV scattered infiltrating cells, black arrowhead: circulating HERV-W ENV positive cells, black dotted line: sub-epithelial HERV-W ENV positive cells, red dotted line: HERV-W ENV positive aggregate clots. A-C and G-J: Bars = 100 µm; D-F’: Bars = 250 µm.

In COVID-19 heart tissue samples (Figure 7), HERV-W ENV was mainly found in endothelial cells from numerous small blood vessels, within cardiac muscle (Figure 7A and 7B) and in the pericardial fatty tissue (Figure 7D and 7E). Endothelial nature of HERV-W ENV positive cells was confirmed with CD31 staining in similar vessel structures from neighboring slides (Figure 7C and 7F). As for each type of tissue, the specificity of the anti-HERV-W ENV monoclonal antibody was confirmed on sections from the same samples showing no staining with an isotype control (Figure 7 G-J). SARS-CoV-2 antigen was not detected in cardiac tissue from studied samples (not shown).

**Figure 7:**
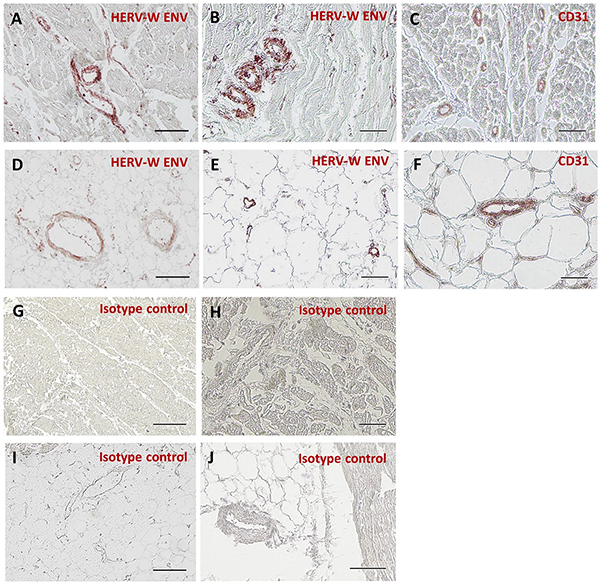
Immunohistological detection of SARS-CoV-2 and HERV-W ENV antigen in post-mortem cardiac tissue from acute COVID-19 cases. Anti-HERV-W ENV (GN_mAb_Env01) murine IgG1 (A, B and D, E), anti-CD31 (C, F) and murine IgG1 isotype control (G-J) antibodies were used on sections of post-mortem heart tissue from COVID-19 patients. Two types of tissues were identified on sections: cardiac muscle (A-C, G, H) and pericardia fatty tissue (D-F, I, J). Murine IgG1 isotype control (G-J) was used to assess possible unspecific background. HERV-W ENV staining (brown/red staining) was analyzed in cardiac muscle (A, B) or pericardial fatty tissue (D, E). In order to identify the HERV-W ENV producing cell type, a CD31 immunostaining (brown/red staining), specific marker of endothelial cells, was performed (C, F). Bars = 100µm

To further address the expression of HERV-W ENV in the central nervous system (CNS) of COVID-19 patients, as previously observed in multiple sclerosis lesions [46], we analyzed sections from tissue samples taken across the cribriform plate, comprising areas of the olfactory bulb and of the nasal mucosa (Figure 8A). This anatomical region was chosen since suggested to be a most probable route of coronavirus passage to the brainstem via olfactory nerve roots within nasal mucosa [75] and since frequently reported anosmia in COVID-19 patients indicates pathological involvement of the olfactory bulb parenchyma [76, 77]. Sections from samples with the upper anatomical brain region, i.e. the frontal lobe, were also made available.

**Figure 8:**
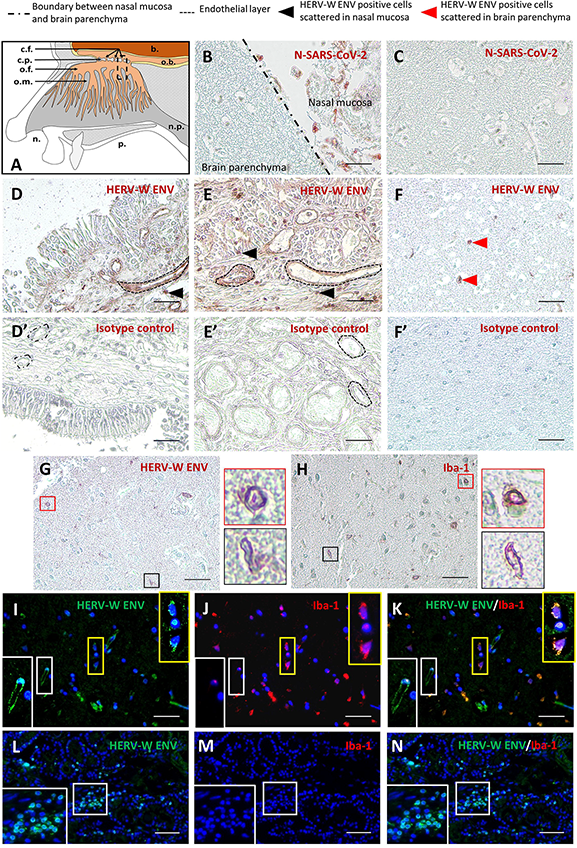
Immunohistological detection of SARS-CoV-2 and HERV-W ENV antigen in post-mortem brain parenchyma of the olfactory bulb and in nasal mucosa tissue from acute COVID-19 cases. (A) Schematic representation of nasal cavity and location of tissue sampling. b. : brain; o.b. : olfactory bulb; n.p. : naso-pharynx; p. : palate; n. : nostrils; o.m. : olfactory mucosa; o.f. : olfactory fibers; c.p. : cribriform plate (ethmoid bone); c.f. : cribriform foramina. Three types of tissues were identified on slides: brain parenchyma (B, C, F, F’, G, H and I-K), nasal epithelium (D, D’) and nasal mucosa (B, E, E’ and L-N). The black dotted line on picture B represents the boundary between brain parenchyma and nasal mucosa. Anti-N-SARS-CoV-2 antibody (B, C), anti-HERV-W ENV (GN_mAb_Env01) murine IgG1 antibody (D-F, G, I and L) and a murine IgG1 isotype control (D’-F’) were used at the same concentration (10 µg/mL). Anti-Iba-1 (H, J and M) antibodies were used for IHC (brown/red staining) or IF (fluorescent staining) on tissue sections at the olfactory brain/nasal mucosa interface or at the upper level within the frontal lobe parenchyma. Detection of SARS-CoV-2 virus antigen in tissues was assessed using an anti-SARS-CoV-2 nucleocapsid antibody (B, C). HERV-W ENV IHC staining was analyzed in nasal epithelium (D), nasal mucosa (E) and brain parenchyma (F, G). Murine IgG1 isotype control (D’-F’) was used to assess possible unspecific background. In order to determine whether microglial cells represent the HERV-W ENV producing cell type, anti-HERV-W ENV (G) and anti-microglia specific marker Iba-1 (H) staining in adjacent sections of brain parenchyma was performed. The morphology of HERV-W ENV positive cells is highlighted with higher magnification of small round cells (G, red square) and small elongated cells (G, black square). Higher magnifications of Iba-1 positive small round (H, red square) and small elongated cells (H, black square) are also boxed. To ascertain that microglia is effectively producing HERV-W ENV, a double IF was performed on a slide with olfactory brain parenchyma and nasal mucosa: HERV-W ENV (I, L, green label) and Iba-1 (J, M, red label). Merged pictures (K, N) illustrate HERV-W ENV/Iba-1 double positive cells (higher magnification: I-K, yellow square). HERV-W ENV positive / Iba-1 negative cells in brain parenchyma (higher magnification: I-K, white square) and nasal mucosa (higher magnification: L-N, white square) are also presented. DAPI was used to stain nuclei (I-N blue staining). Black dotted line: endothelial layer, black arrowhead : HERV-W ENV positive cells scattered in nasal mucosa, red arrowhead : HERV-W ENV positive cells scattered in brain parenchyma. b. : brain; o.b. : olfactory bulb; n.p. : naso-pharynx; p. : palate; n. : nostrils; o.m. : olfactory mucosa; o.f. : olfactory fibers; c.p. : cribriform plate (ethmoid bone); c.f. : cribriform foramina. Bars = 100µm.

Immunohistology results for both SARS-CoV-2 and HERV-W ENV antigens in brain parenchyma or nasal mucosa areas are presented Figure 8. In sections at the CNS-nasal tissue interface presented in Figure 8B, a strong SARS-CoV-2 N antigen staining indicating viral replication was seen in nasal mucosa but not in neighboring CNS areas of the olfactory bulb. SARS-CoV-2 antigen was not detected in various areas of CNS sections within olfactory bulb or frontal brain parenchyma, as illustrated in Figure 8C.

A strong HERV-W ENV staining was seen in nasal mucosa, involving infiltrated lymphoid cells and, as already seen in pulmonary and cardiac tissues, the endothelium of blood vessels (Figure 8D and 8E). Conversely to SARS-CoV-2 antigen, HERV-ENV was also detected in the olfactory bulb parenchyma in glial cells (Figure 8F), which revealed to have the morphology of microglia and were stained by a microglial maker, Iba-1 (Figure 8G and 8H). Higher magnifications of HERV-W ENV positive microglial cells within brain parenchyma are boxed and presented in Figure 8G and 8H. The absence of staining with the isotype control further confirmed the specificity of previous staining in COVID-19 tissue samples (Figure 8 D’-F’). To further confirm the expression of HERV-W ENV in microglia with COVID-19 brain parenchyma, a double immunostaining was performed with the fluorophore-labeled same monoclonal antibodies against HERV-W ENV and Iba-1. Microglia-like cells positive for HERV-W ENV revealed to be co-stained with the antibody against Iba-1, and could thus be confirmed to be microglial cells (Figure 8 I-K, boxed in yellow with higher magnification on the right side). On the same pictures, endothelial cells from an HERV-W ENV positive blood vessel wall were consistently negative for Iba-1 detection (Figure 8 I-K, boxed in white with higher magnification on the left side). In parallel, sections from nasal mucosa showed staining of infiltrated lymphoid cells also consistently negative for Iba-1 (Figure 8 L-N, boxed in white with higher magnification on the left side). Clinical data and summary of IHC results from present tissue sections are respectively presented in **Supplementary Tables S6 and S7**.

## Discussion

The COVID-19 pandemic is threatening populations worldwide because of severe forms requiring well-equipped hospitals with sufficient capacity of intensive care. Vaccination has proved to be efficient despite a lack of knowledge on the long-term protective immunity and on the possible emergence of variants no longer under control with present vaccines. However, this new disease revealed how heterogeneous and various the clinical profiles or the evolution of acute and post-acute COVID-19 may be, with syndromes or symptoms not directly related to the viral infection.

In the present study we have analyzed the induction of HERV expression during COVID-19 disease. Firstly, *in vitro* results showed that SARS-CoV-2 can trigger both HERV-W and HERV-K *ENV* RNA transcription, in short-term PBMC cultures from healthy individuals, after a single exposure to SARS-CoV-2 virus. In contrast to the induced HERV-W and -K transcription, only HERV-W ENV protein was detected during short-term primary cultures of PBMC from about 30% of healthy donors.

As wild-type SARS-CoV-2 spike protein trimer induced the production of IL-6 from PBMC of all donors, either responding or non-responding with HERV activation, IL-6 cannot be responsible for the induction of HERV-W ENV expression. The observed HERV-W activation is therefore not likely to be induced by cytokines or by inflammation due to viral infection, but by SARS-CoV-2 spike protein itself, as further shown. Cytofluorometry analysis confirmed that HERV-W ENV protein expression was predominantly and strongly induced in CD3^low^ T-lymphocytes within the CD3^+^ T-cell population. T-lymphocytes that underwent a very recent contact with antigenic components of SARS-CoV-2 may dynamically become activated CD3^+^ T-cells as previously described [72]. Alternatively, in COVID-19 patients, our observations may corroborate previous reports describing superantigen motifs of SARS-CoV-2 spike protein [73], and may involve cellular mechanisms associated to lymphopenia and hyperinflammation as already characterized for other emerging viruses (e.g., Ebola or Lassa) [78, 79]. However, because HERV-W ENV has also been shown to display superantigen-like effect [51], the origin of this short-term effect on T-lymphocytes may be questioned. Results from a recent study potentially provide a first answer, since showing correlated expression between HERV-W ENV and markers of exhaustion in T-lymphocytes from severe COVID-19 cases [60].

Of note, HERV activation occurs without signs of infection of lymphocytes or monocytes by SARS-CoV-2, while a recombinant trimer of its spike protein without stabilizing mutations [74] appeared sufficient to reproduce similar HERV-W and -K *ENV* RNA stimulation as well as HERV-W ENV protein production in lymphoid cells, albeit in a subset of donors. This type of HERV activation mediated by an interaction between a triggering virus and a specific receptor on certain cells has already been described, e.g., with HHV-6A [33]. Because SARS-CoV-2 induced HERV-W ENV expression in human lymphoid cells that neither express ACE2 nor TMPRSS2 and does not infect them, additional undetermined receptors are expected to mediate HERV activation by SARS-CoV-2, which should now be further studied with recent data on alternative receptors [80].

The study further addressed the frequency of HERV expression in COVID-19 patients. In hospitalized individuals with different severity status, HERV-W ENV protein was confirmed to be expressed at the membrane surface of T-lymphocytes but mainly in CD3low T-cells. A lower but significant detection was also found in B-lymphocytes, which corresponds to a cell type already known to be permissive for HERV-W expression [81, 82]. However, HERV-W expression was not observed in T-cells before its first observation in COVID-19 [60].

After the recent characterization of a soluble hexameric form of HERV-W ENV in MS brain lesions [70], we here observed its presence as a circulating antigen in COVID-19 plasma or sera. In sample series from patients in intensive care unit, a significant correlation was found between HERV-W ENV detection on T-lymphocytes and the soluble antigen in plasma. Though moderate, this correlation was seen between blood cells and plasma devoid of cells, which indicates a relationship between expressing lymphocytes and the release of this soluble antigen in blood. Plasma HERV-W ENV antigenemia was confirmed to be significantly detected in hospitalized patients with more-or-less severe COVID-19 but not in tested healthy controls, whereas HERV-K ENV protein was never detected in plasma or serum. Importantly, HERV-W ENV antigenic levels strongly correlated with disease severity and were significantly different from healthy controls. All severe COVID-19 cases were tested positive for HERV-W ENV protein in plasma, which points to a marker of disease severity as already shown with blood lymphocytes [60]. We also report HERV-W ENV levels in a series of COVID-19 cases that were selected based on positive SARS-CoV-2 PCR, independent of disease severity. Within this series, HERV-W ENV antigenemia was detected in about 20% of samples only, a number quite similar to the percentage of PBMC donors that showed HERV-W ENV positivity in reaction to SARS-CoV-2 challenge *in vitro*. Altogether, these data suggest that a percentage of individuals with an underlying susceptibility to symptomatic and/or severe evolution may be linked to the activation of HERV-W ENV expression. This is also consistent with the significant increase in frequency and in antigenemia with disease severity in the previous series with hospitalized patients (Cf. Figure 5D), similarly to the previously shown increased and predictive expression on T-lymphocytes of such inpatients [60]. In short term PBMC cultures non-responding to SARS-CoV-2 exposure, we had observed HERV RNA levels below the levels of non-exposed controls. Such decreased levels may be explained by the activation of HERV-inhibitory pathways and effectors. Thus, an inter-individual variability in the potency of HERV inhibitory mechanisms, possibly with an (epi) genetic origin, may provide an explanation for non-universal activation of HERV-W ENV upon SARS-CoV-2 challenge. In fact, most PCR positive individuals do not develop major symptomatology after SARS-CoV-2 infection, including about 35% of asymptomatic cases [83].

Furthermore, analyses of other blood parameters showed that the neutrophils counts, reported to be most often increased in COVID-19 with worsening evolution [84, 85], also paralleled HERV-W ENV antigenemia with values above the upper normal limit in all hospitalized patients with ENV positive plasma (Cf. **Supplementary Table S3**). Moreover, since patients from different geographic areas and time periods of the pandemic were analyzed and were confirmed to have been infected by different variants of SARS-CoV-2 (**Supplementary Table S3**), the global results are not expected to be influenced by such variables.

To study HERV-W ENV expression beyond blood cells and plasma or serum, we have analyzed slides from tissue samples obtained at autopsy from patients deceased from acute COVID-19. SARS-CoV-2 N antigen corroborating viral replication was readily detected in epithelial cells within lungs and nasal mucosa but not in studied sections from brain parenchyma, even in olfactory bulb sections neighboring nasal mucosa with noticeable ongoing infection. The present results did not show any infection by SARS-CoV-2 of the central nervous system (CNS) nor of the cardiac tissues in the studied cases. HERV-W ENV was strongly expressed in lymphoid infiltrates in tissues surrounding lung alveola and within nasal mucosa. It was clearly and frequently detected in blood vessels endothelium from all tissues including CNS, which revealed quite numerous within cardiac muscle and pericardiac fatty tissue. In the CNS, HERV-W ENV expression was found in scattered cells that were confirmed to be microglia. Finally, strong HERV-W ENV staining was often detected in aggregated cells corresponding to thrombotic structures in blood vessels from the lung sections. Globally, positive endothelial cells from blood vessels were seen in all studied tissues and all HERV-W ENV expressing cells did not correspond to phenotypes seen to be infected with SARS-CoV-2. This discrepancy is well illustrated by infected lung epithelial cells without HERV-W ENV expression and non-infected alveolar macrophages presenting strong HERV-W ENV membrane staining within the same lung section. This is consistent with our observations of HERV-W ENV protein presence on lymphocytes without infection by SARS-CoV-2 in COVID-19 patients and in PBMC, after *in vitro* activation with infectious SARS-CoV-2. Most importantly, it shows HERV-W ENV expression in tissue-infiltrated lymphocytes and macrophages within affected organs, like in blood of COVID-19 patients. Therefore, results from immunohistochemistry analyses indicate that HERV-W ENV expression is intimately associated with organs and cells involved in COVID-19 associated or superimposed pathology beyond tissue inflammation mediated by immune cells or cytokines, e.g., in vasculitis or intravascular thrombotic processes [29, 55]. Moreover, given known HERV-W involvement in MS pathogenesis [43, 46, 48] or in certain psychoses associated with inflammatory biomarkers [58, 86], the presently observed HERV-W ENV expression in microglia strongly suggests a role in neurological symptoms and cognitive impairment mostly occurring or persisting during the post-acute COVID-19 period [14, 17, 64, 66, 77].

In acute primary infection, one must also consider the pathogenic effects on immune cells resulting in an hyperactivation of the innate immunity via HERV-W ENV-mediated TLR4 activation [53] with a possible contribution to the frequently observed lymphopenia along with an adaptive immune defect. The induction of autoimmune manifestations [87–90] as previously shown to be induced with HERV-W ENV (previously named MSRV) in a humanized mouse model [47], should also be considered. Altogether, these data indicate that HERV-W ENV does not simply represent a biomarker of COVID-19 severity or evolution, but is also likely to be a pathogenic player contributing to the disease severity.

To date, various studies have considered many parameters in COVID-19 patients, but none has addressed HERV activation and expression of HERV proteins initiating and perpetuating pathogenic pathways underlying worsening clinical evolution and long-term pathology as seen with the now emerging post-COVID “wave”. The latter represents millions of patients suffering from various and numerous symptoms, who develop long-term disabling active pathology for which no rationalized understanding nor therapeutic perspective can be proposed to date.

In face of this challenging situation, data from the present study altogether call for evaluating HERV-W ENV as a marker of severity but also as a potential therapeutic target in COVID-19 associated syndromes. This should lead to already existing therapeutic molecules targeting HERV-W ENV in clinics (ClinicalTrials.gov Identifier: NCT04480307). In addition, further characterization of HERV-W expression in population should help understanding of the individual differences in the response to SARS-CoV-2 infection, and may provide the critical set up for the identification of novel biomarkers specific for severe forms of COVID-19 and/or for long-term evolution of some post-COVID symptoms.

## Data Availability

All data produced in the present work are contained in the manuscript

## Acknowledgments

We acknowledge World Reference Center for Emerging Viruses and Arboviruses (WRCEVA) and UTMB investigator, Dr. Pei Yong Shi for kindly providing recombinant NeonGreen virus based on 2019-nCoV/USA_WA1/2020 isolate and VirPath from the CIRI, Centre International de Recherche en Infectiologie, Lyon, France (B. Lina, A. Pizzorno, O. Terrier and M. Rosa-Calatrava), for providing us with the BetaCoV/France/IDF0571/2020 virus. We also acknowledge Dr. Vinca Icard responsible for the BioCovid collection at the “Hospices Civils de Lyon (HCL)” and the “Centre de ressources Biologiques (CRB)” of the HCL such as Healthy and Covid-19 donor’s samples supplier. We acknowledge the contribution of the cytometry CELPHEDI / AniRa facility depending of SFR Biosciences (UAR3444/CNRS, US8/Inserm, ENS de Lyon, UCBL) and more particularly Sébastien Dussurgey for his help for cytofluorometry analyzes. We acknowledge Dr. Sandrine Levet for her kind participation for manuscript revision. This work was supported by ANR-CoronaPepStop (ANR-20-COVI-000) and Fondation de France to BH and Agence Nationale de la Recherche (ANR, France)-COVERI to HP.

HP, BC, JB, JP and NQ receive compensation from Geneuro-Innovation for their work. PK received consulting fees from Geneuro SA. Other co-authors do not declare conflict of interest.

**Tables S1 to S7 and Figures S1 to S6 are presented with legends in supplementary material.**

## Supplementary Table and Figures

**Supplementary Table S1. RT-qPCR HERV-W *ENV* and HERV-K *ENV* results from cultured PBMC with or without exposure to infectious SARS-CoV-2.**

Mean 2^-ΔΔCt^ and standard deviation (SD) of triplicate results from RNA of PBMC cultures from 11 donors from #26 to 43, in presence or absence of SARS-CoV-2 infectious virus. RNA was extracted from 3 cultured wells and tested in triplicate at 2h or, for kinetics analysis represented in Figure 1, from a single well at each time point (donors #26-29). RT-qPCR protocol with B2M reference RNA was performed in duplicate measures on each cDNA according to Material and Methods. PBMC cultures from donors presented for immunofluorescence or cytofluorometry analyses could not all be used to extract RNA in parallel, due to the limited volume of blood donations, and therefore, to the limited number of available cells.

**Supplementary Table S2. Clinical and biological data (Lyon_2)**

A. Bioclinical data
B. Biological parameters

**Supplementary Table S3. Clinical and biological data (Lyon_3)**

A. Bioclinical data
B. Biological parameters

**Supplementary Table S4. Clinical and biological data (Zaragoza)**

A. Bioclinical data
B. Biological parameters

**Supplementary Table S5. Origin, description and reference of post-mortem samples for the IHC study**

**Supplementary Table S6: clinical data from COVID-19 samples for immunohistochemistry (post-mortem tissues from COVID-19 patients)**

**Supplementary Table S7. Global results from IHC analyses on series of slides from COVID-19 post-mortem tissue samples.**

**Supplementary Figure S1: Kinetics of HERV-W *ENV*, HERV-K *ENV* and SARS-CoV-2 *N* RNA detection in PBMC from healthy blood donors exposed to SARS-CoV-2.**

(A-B) PBMCs from 4 healthy blood donors were exposed to SARS-CoV-2 virus (MOI: 0.1) in parallel to mock-control (corresponding to the absence of virus) and HERV-W *ENV* (A) and HERV-K *ENV* (B) mRNA levels were assessed at 2h, 19h and 24h post-inoculation (pi) by RT-qPCR using specific primers (Table 1). Results are presented as the fold change of the corresponding non-infected condition. (C) The mRNA levels of SARS-CoV-2 *N* mRNA of PBMCs cultures from the same 4 healthy blood donors, exposed (plain histograms) or not (hatched histograms) to infectious SARS-CoV-2 (MOI:0.1), were analyzed by RT-qPCR. Results are presented as RT-qPCR fold change compared to the corresponding non-infected condition. NI: Control culture Not Inoculated with SARS-CoV-2 virus; pi: post-inoculation. The quantity of SARS-CoV-2 *N* mRNA was monitored 2, 19 and 24 hours after SARS-CoV-2 infection (black histograms). The quantity of SARS-CoV-2 *N* mRNA corresponds to the initial input of virus and did not increase with time but significantly decreased, thereby indicating the absence of productive SARS-CoV-2 replication in PBMCs cultures.

**Supplementary Figure S2: Immunofluorescence and RT-qPCR specificity controls.**

Simian Vero cells (African Green Monkey) are routinely used for the preparation of viral stocks. Non-infected Vero cells were used as negative controls for immunostainings (A, C) while SARS-CoV-2 infected Vero cells (B, D) were used as positive controls for anti-N- and anti-S-SARS-CoV-2 immunodetection (respectively B and D, red labelling). Vero (non-human) cells showed no expression of HERV-W *ENV* and HERV-K *ENV* mRNA at 2h pi and 24h pi (respectively E and F) nor of HERV-W ENV protein (A-D, green labelling). The efficiency of viral replication was monitored between 2h post-infection (pi) and 24h pi by RT-qPCR using *N*-SARS-CoV-2 primers (G). RT-qPCR results were presented as the fold change compared to the corresponding non-infected condition (NI).

(H-N) HEK293T cells were transfected with plasmids encoding for *GFP*, HERV-W *ENV*, *ERVWE1* (syncytin-1) or HERV-K *ENV*. GN_mAb_Env01 (anti-HERV-W ENV) and GN_mAb_EnvK01 (anti-HERV-K ENV) do not stain any GFP expressing cells (H, L, red staining). GFP transfected cells allowed to determine the transfection efficacy, evaluated around 80%. GN_mAb_Env01 detects HERV-W ENV on transfected cells (I, green staining) but did not detect the homologous Syncytin-1, forming syncytia in transfected cells cultures (J, syncytia induced by syncytine-1 expression are delineated by dotted lines). Anti-HERV-K ENV was also very efficient to detect its target antigen in HERV-K ENV transfected cells culture (K). Background signal of secondary antibodies was assessed in the absence of primary antibodies (M, N).

In order to evaluate the background level generated by the secondary antibodies, negative controls in the absence of primary antibodies controls were made with PBMC cultures from 3 healthy donors (donors # 7-9), exposed for 72 hours to the infectious SARS-CoV-2 virus inoculated at MOI:0.1 (R-T), or sham-inoculated (O-Q). IF controls without primary antibodies are also presented for Vero cells infected with SARS-CoV-2 at MOI:0.1 for 24 h (U). DAPI was used to stain nuclei (A-D and H-U, blue staining). Bars : A-D and H-N = 50µm; O-U = 100µm.

**Supplementary Figure S3: Expression of *ACE2* in PBMC from healthy individuals and cell viability assay.**

(A) RT-qPCR with specific *ACE2* primers in PBMCs from 11 healthy donors, showing the absence of detectable mRNA expression in human PBMC. (B) PBMCs isolated from 5 healthy blood donors (donors # 17 to 21), were treated, or not (NT), with 0.5 µg/mL or 2.5 µg/mL of active trimer Spike recombinant protein. Cell viability in cultures was assessed using the Promega® “CellTiter-Glo 2.0®” kit assay. Results were expressed as the percentage of viability measured in the non-treated culture at 24 h post treatment (hpt). Statistical analysis: Sidak’s multiple comparison. 2 types of comparisons were performed: comparison between NT and treated condition and comparison between 24 hpt and 48 hpt conditions. (n.s: *p*>0.5; *: 0.5<*p*<0.01).

**Supplementary Figure S4: Examples illustrating the gating strategy for the cytofluorometry analysis of PBMC from COVID-19 patients and controls.**

Dot plots of a representative HBD (A-B, left panel) and a COVID-19 patient (C,D, right panel). Analysis of the percentage of HERV-W ENV positive cells in CD14 and CD3 T cells (after the suppression of CD14 cells from the gating), with identification of CD3^high^ and CD3^low^ T cell subpopulations (A, C). Analysis of the percentage of HERV-W ENV positive cells in CD19^+^ B cells population (B,D). Of note, the specific identification of CD3^low^ T cells with an increased size was confirmed with the suppression of CD14 positive monocyte cells from the gating. The analysis was done by acquiring at least 50 000 events in the PBMC gate.

**Supplementary Figure S5. Absence of detection of HERV-K ENV in plasma of COVID-19 patients and of correlation between HERV-W ENV and SARS-CoV-2 serology.**

(A) Scatter plot of quantification of soluble HERV-W ENV (Y axis; HERV-W ENV S/N ratio) and quantification of SARS-CoV-2 antigens (X axis) in plasma of Lyon_3 cohort. Spearman’s table was used to determine the correlation coefficient. (B-D) Immunodetection of HERV-K ENV in 11 plasma from HBD (blue dots) and 21 plasma from COVID-19 patients from Lyon (France, “Lyon_3 cohort”; (red dots) after protein extraction and Wes analysis under denaturing conditions with primary antibody GN_mAb_EnvK01. Electrophoregrams (B) and digital western blots (C) of a representative HBD (blue panel) and a representative COVID-19 patient (red panel). The AUC of the target hexamer HERV-K ENV (electrophoregram peak about 60-90 kDa) was measured and expressed as HERV-K ENV S/N ratios (D). S/N ratio below the positivity threshold (< 1) were corrected to a value equal to 1.

**Supplementary Figure S6. Specificity controls for IHC analyses.**

IHC of HERV-W ENV using GN_mAb_Env01 (A,B,E,F,I,J) and anti-N-SARS-CoV-2 (C,D,G,H,K,L) antibodies were performed on non-COVID-19 lung tissue sections of normal appearing tissue from 3 lung necropsies from non-COVID-19 patients (lung cancer) (A-D, E-H, I-L). Bars = 250µm

