## Supplementary figures and images for "SARS-CoV-2 induces human endogenous retrovirus type W envelope protein expression in blood lymphocytes and in tissues of COVID-19 patients"

### Supplementary Figure S1

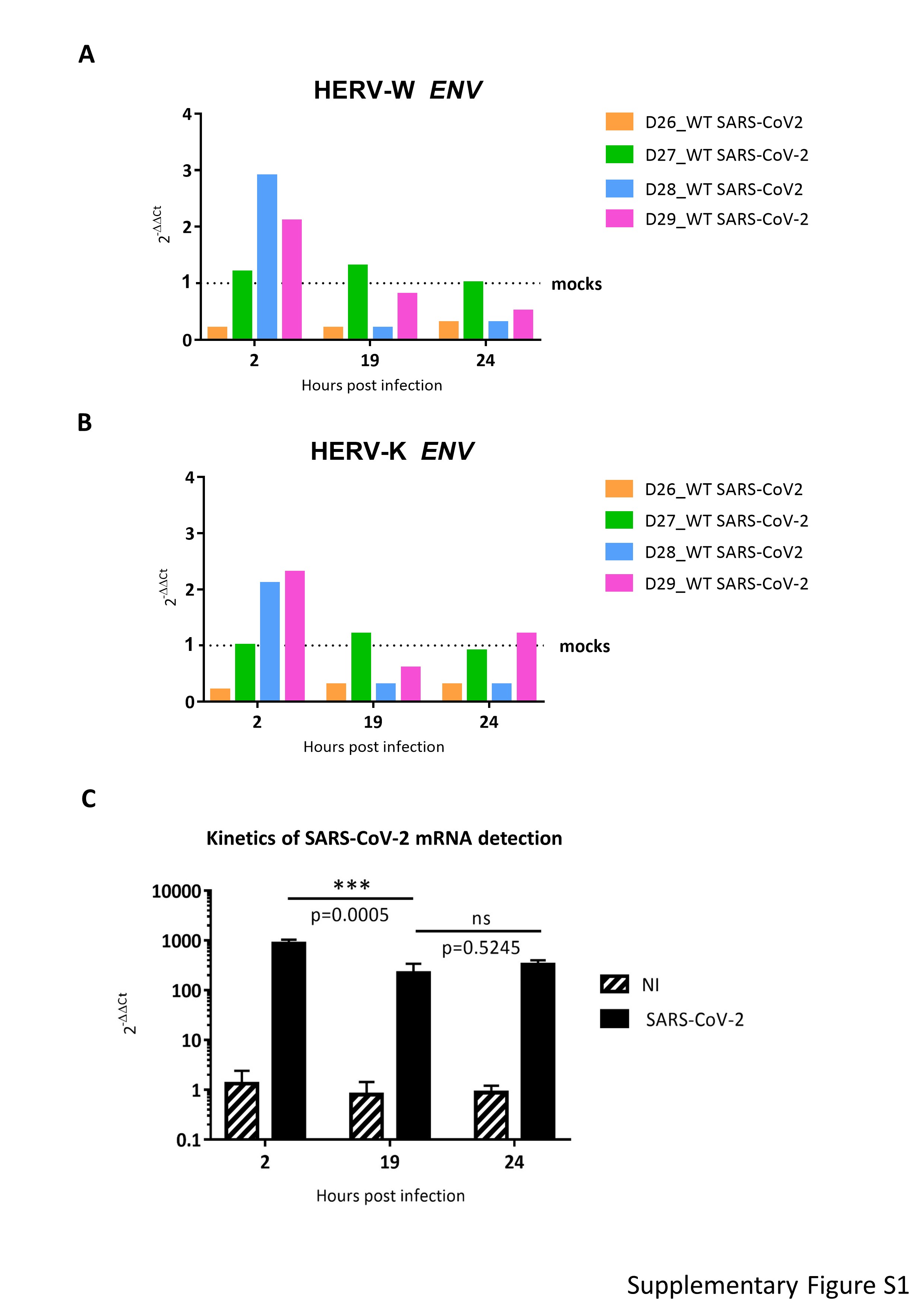

### Supplementary Figure S2

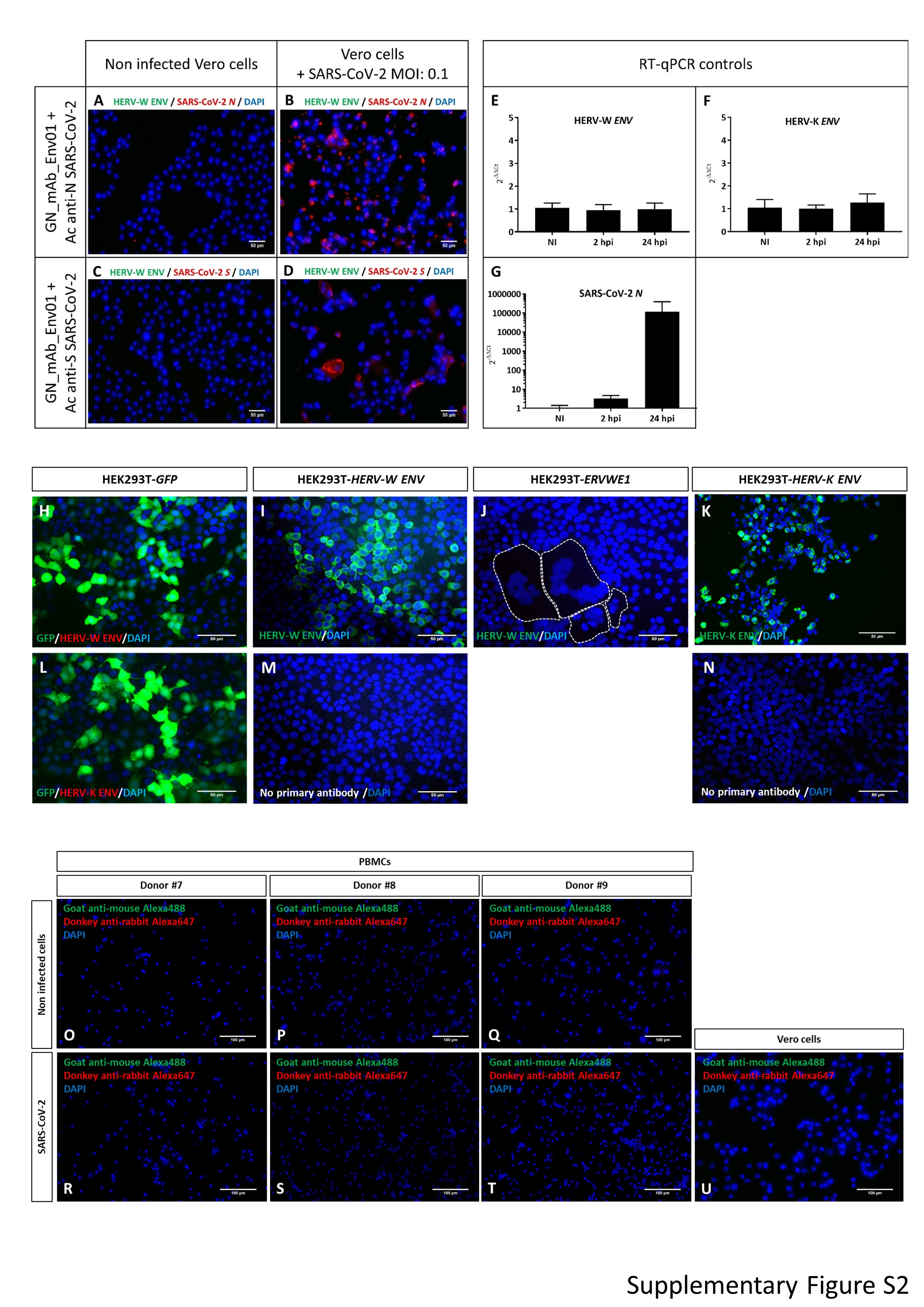

### Supplementary Figure S3

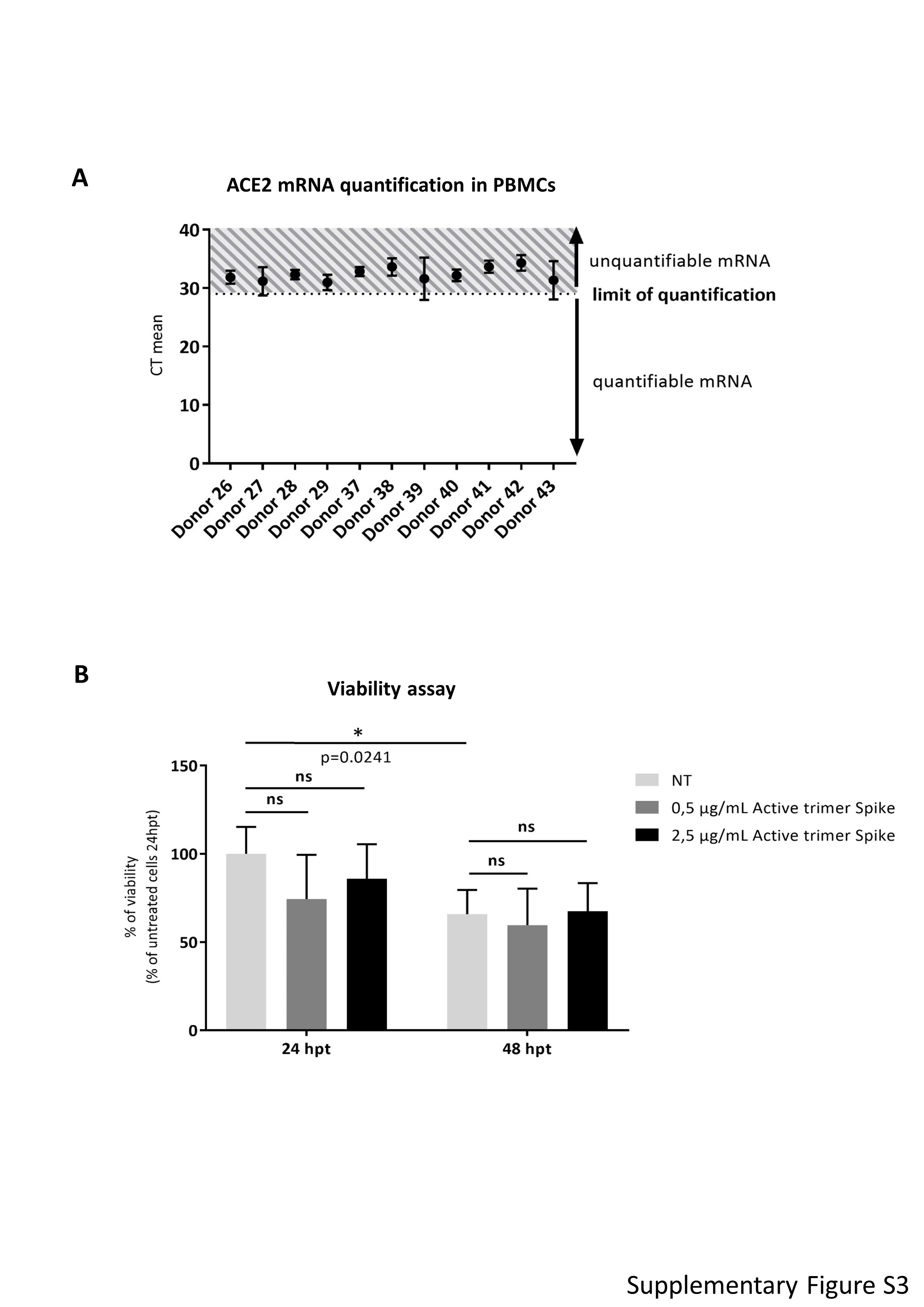

### Supplementary Figure S4

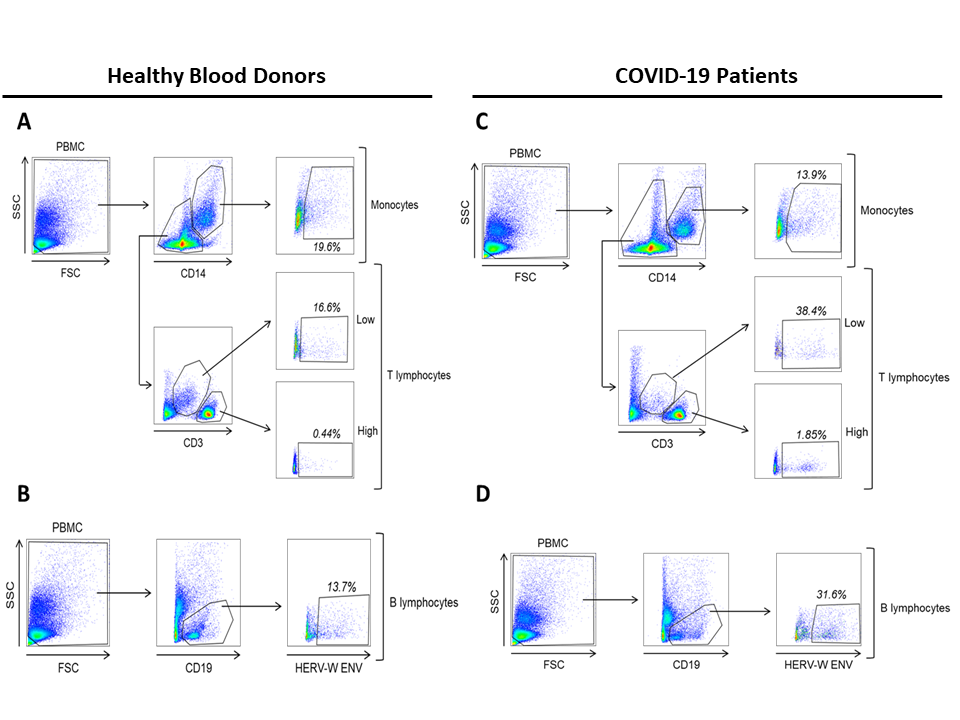

### Supplementary Figure S5

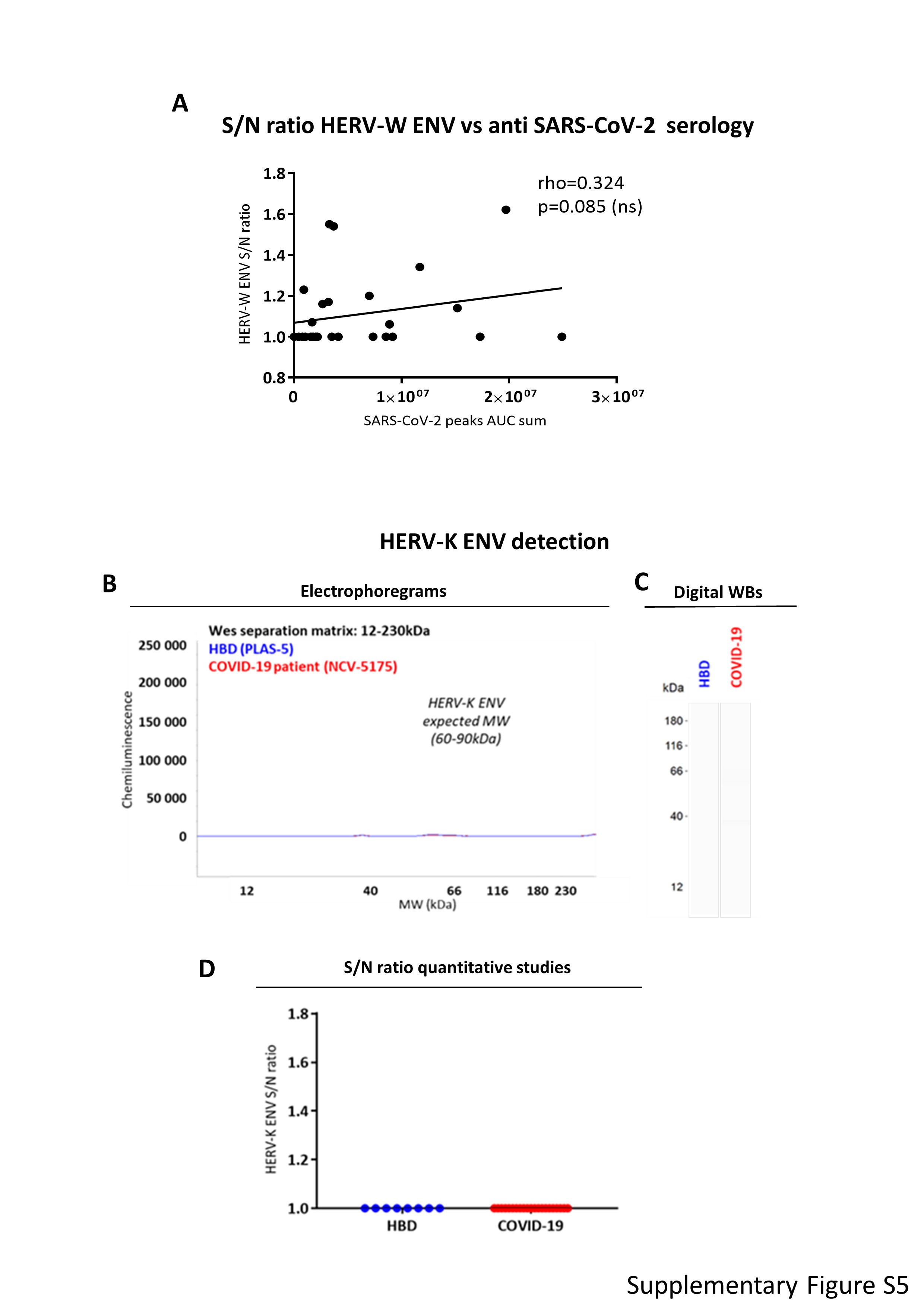

### Supplementary Figure S6

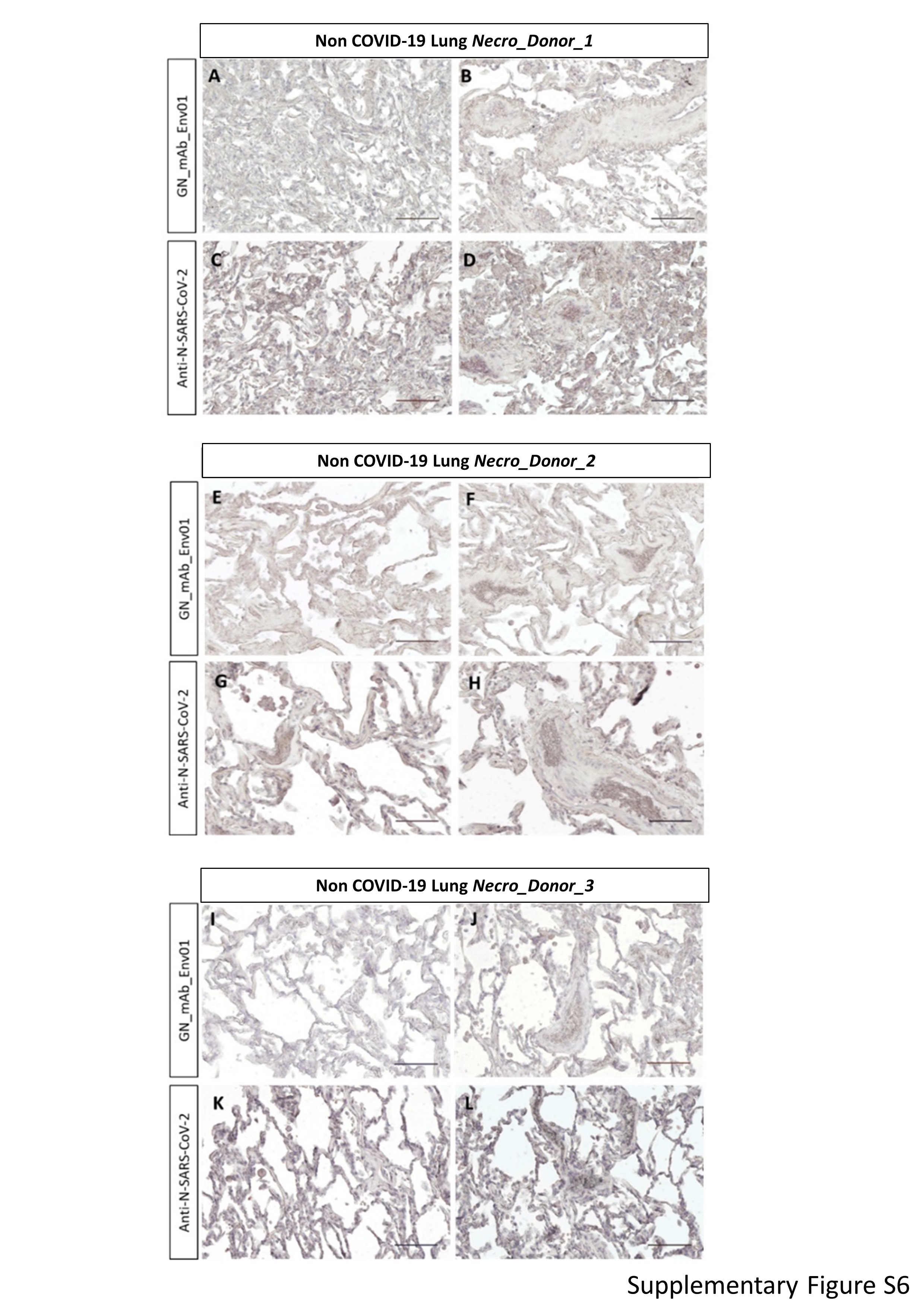

### Supplementary Table S1

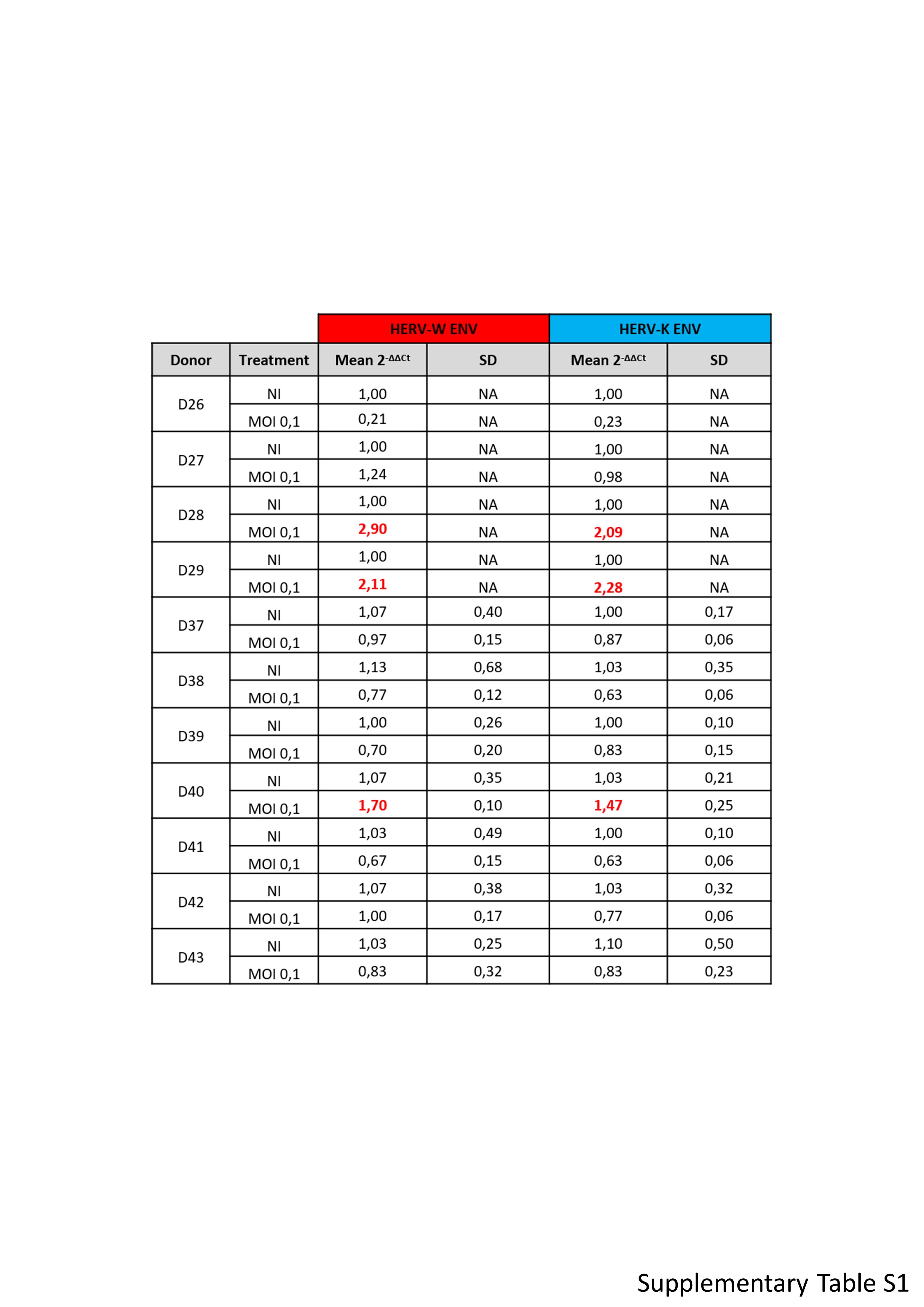

### Supplementary Table S2

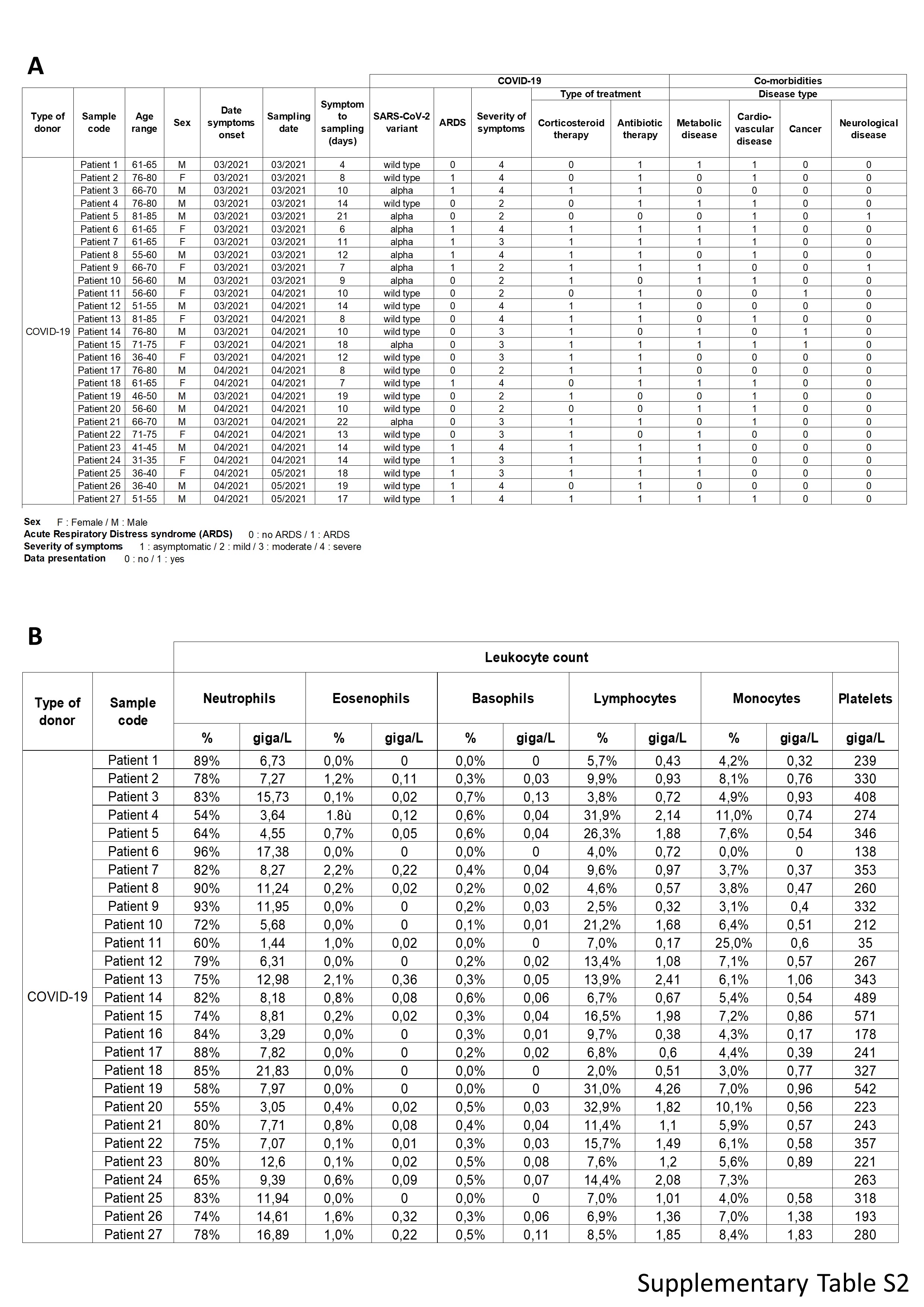

### Supplementary Table S3

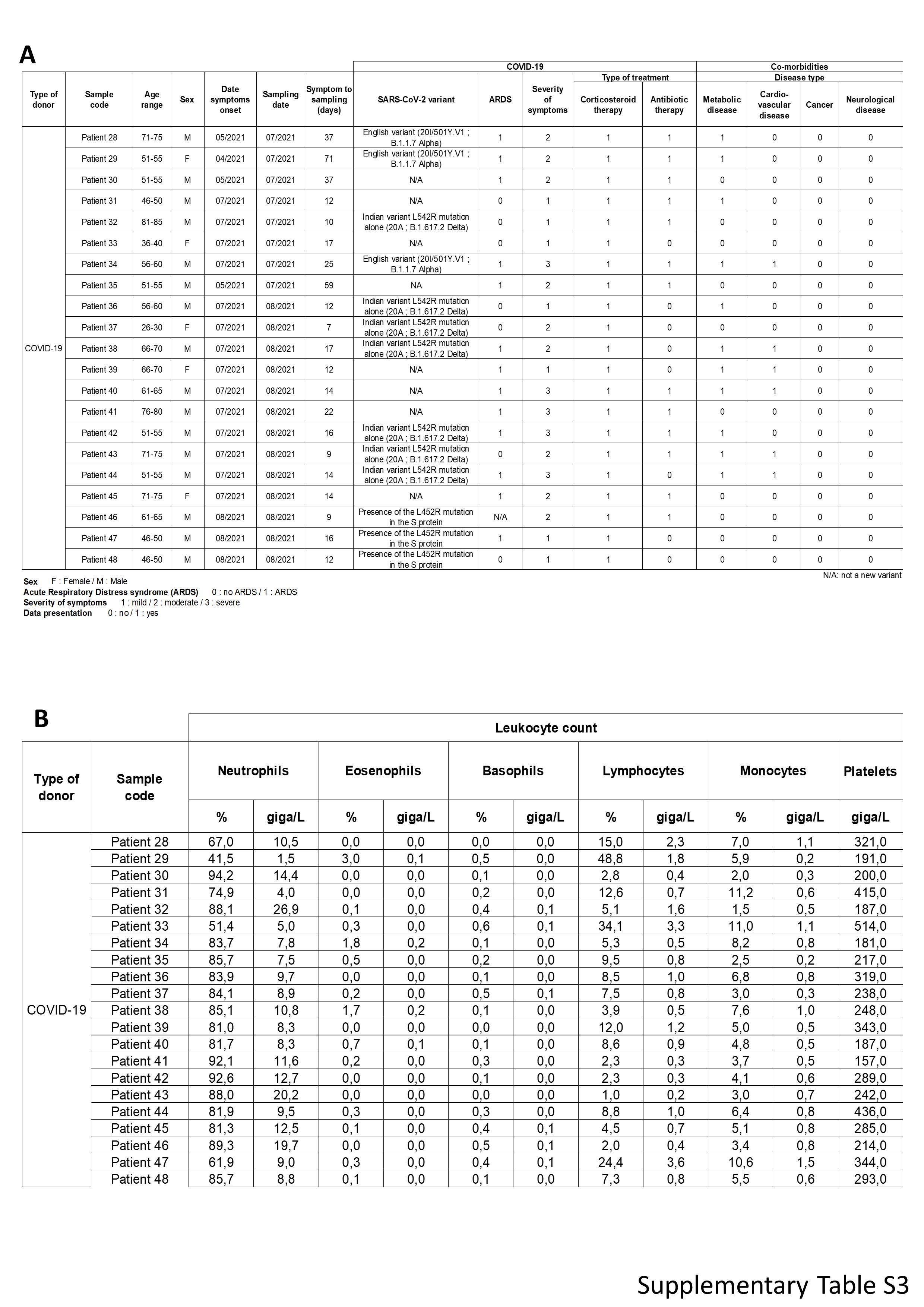

### Supplementary Table S4 A

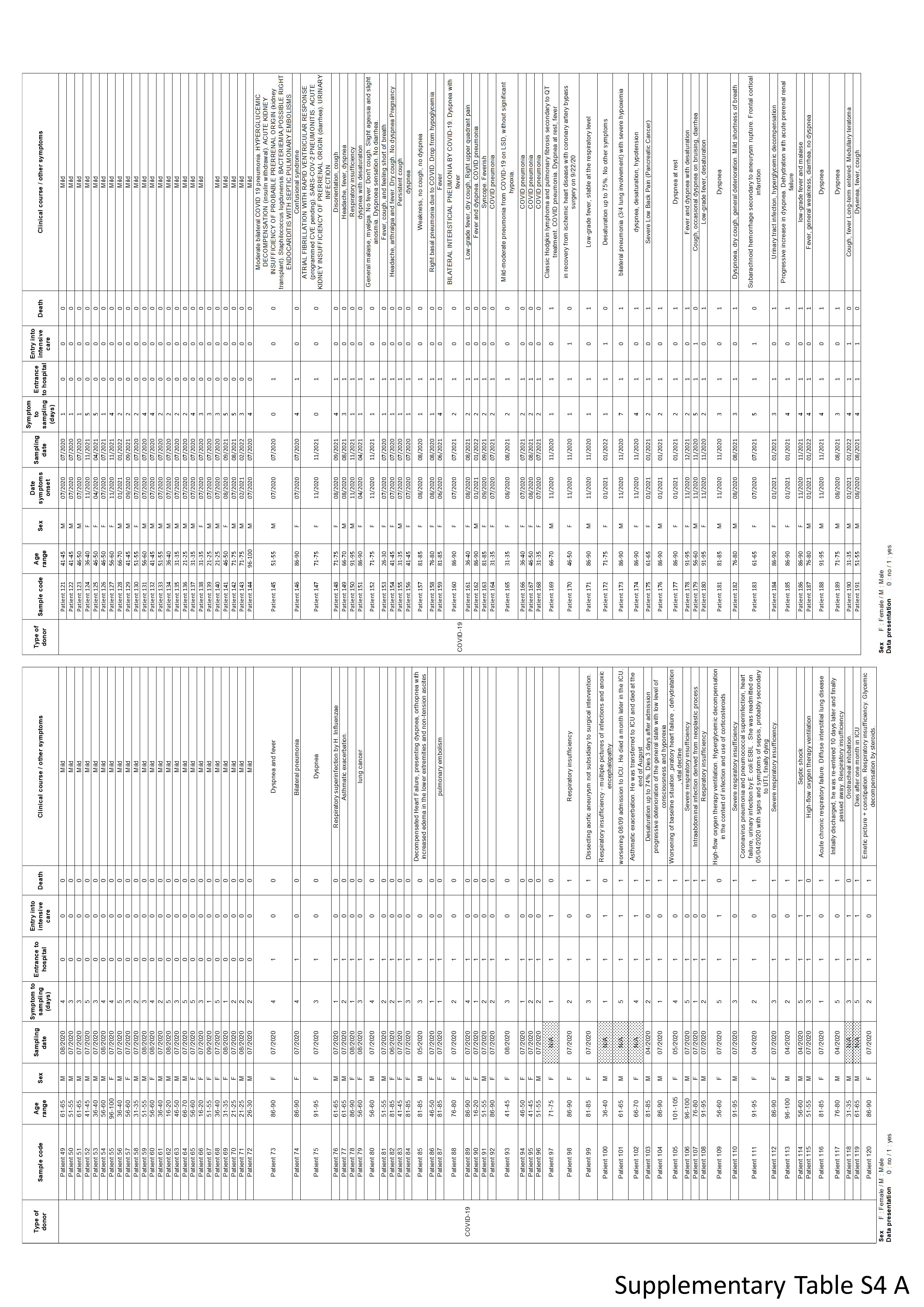

### Supplementary Table S4 B

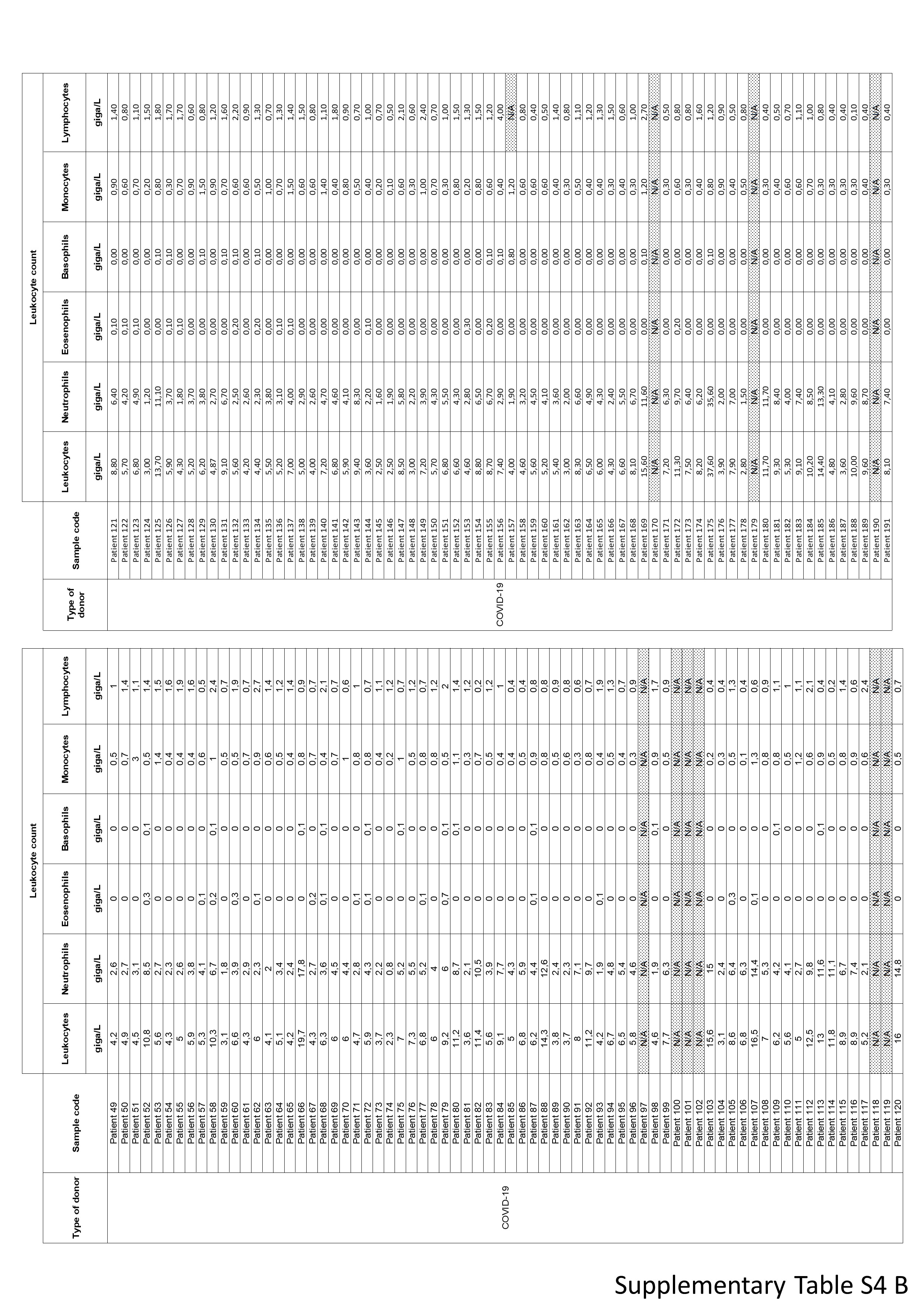

### Supplementary Table S5

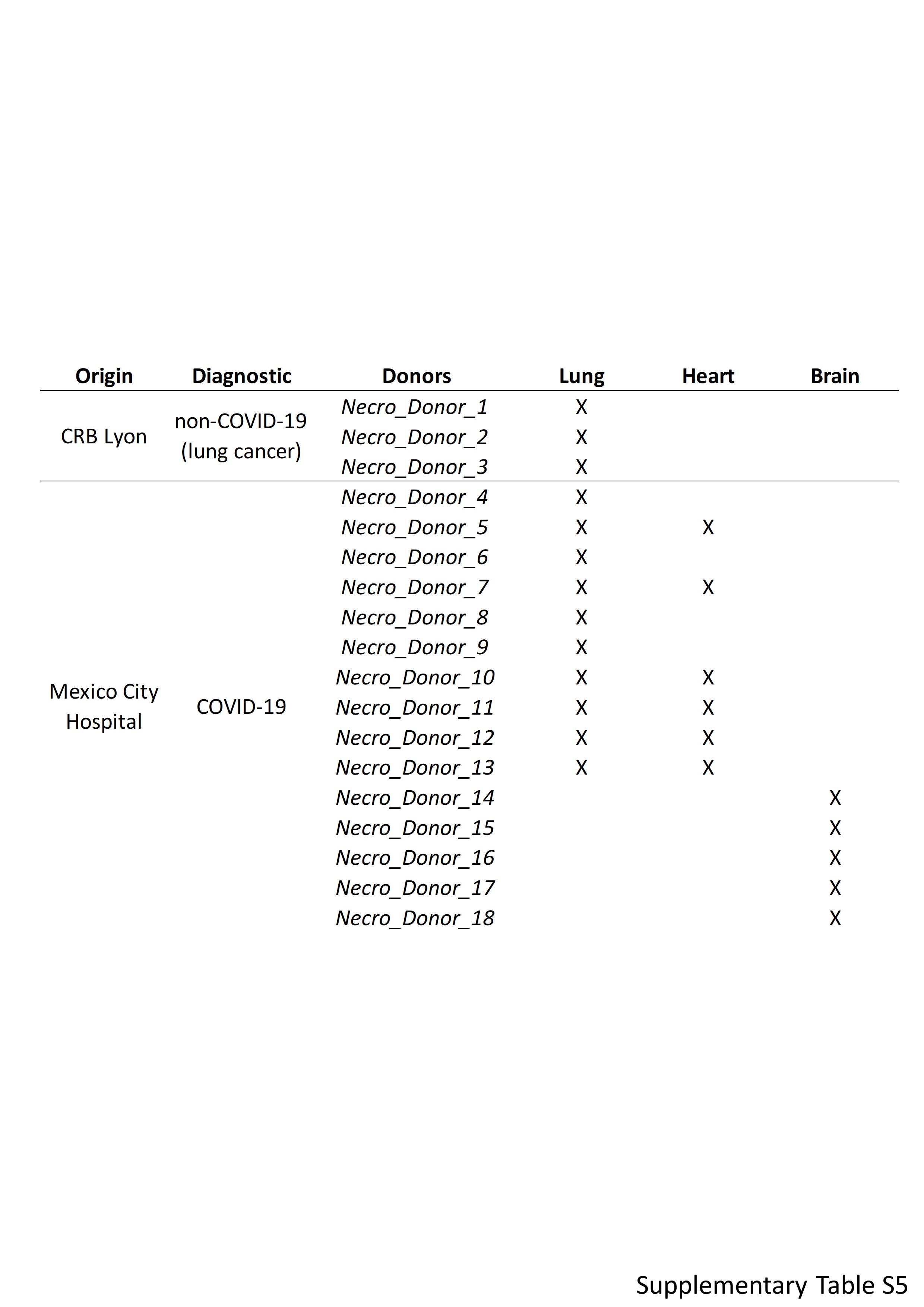

### Supplementary Table S6

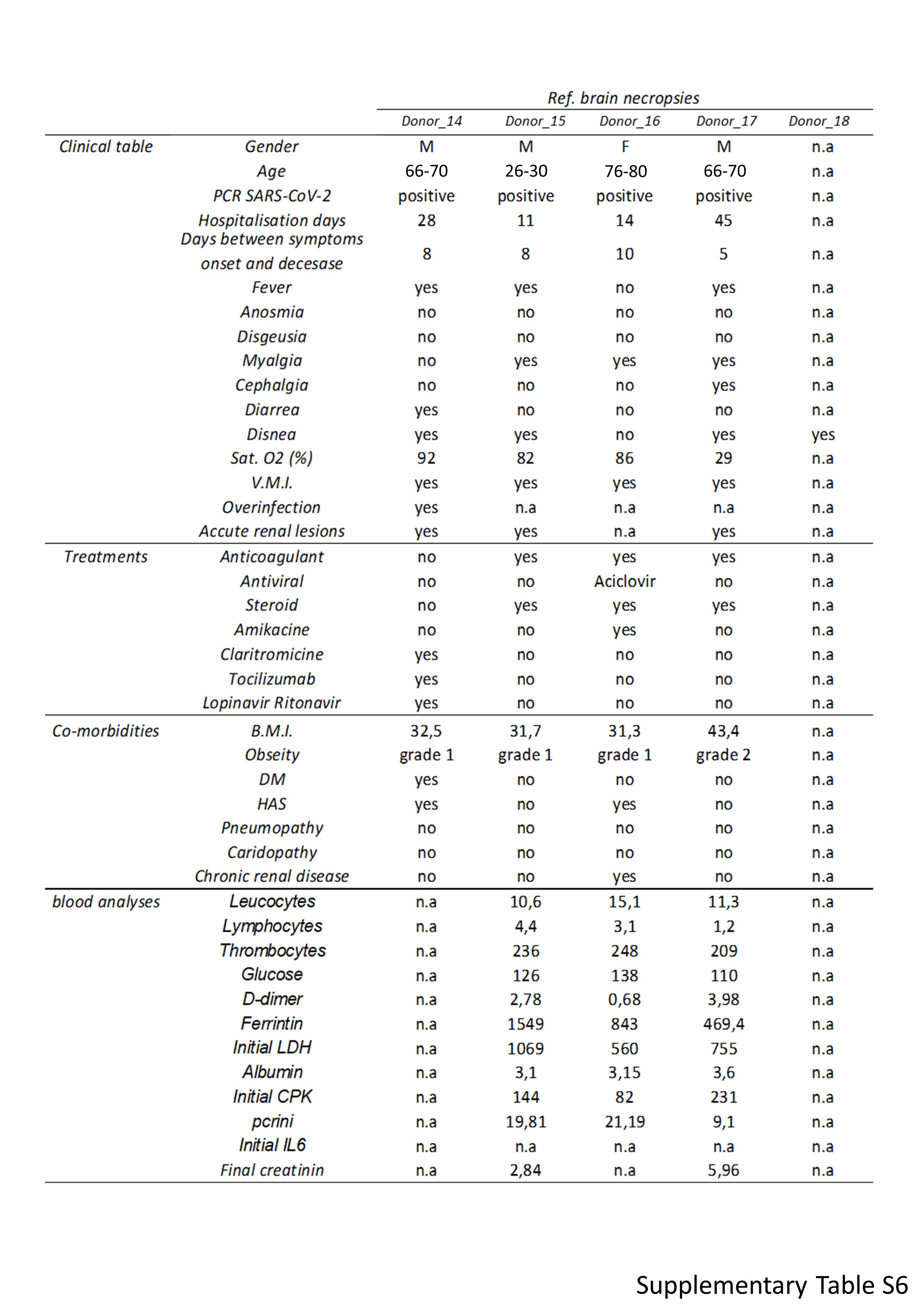

### Supplementary Table S7

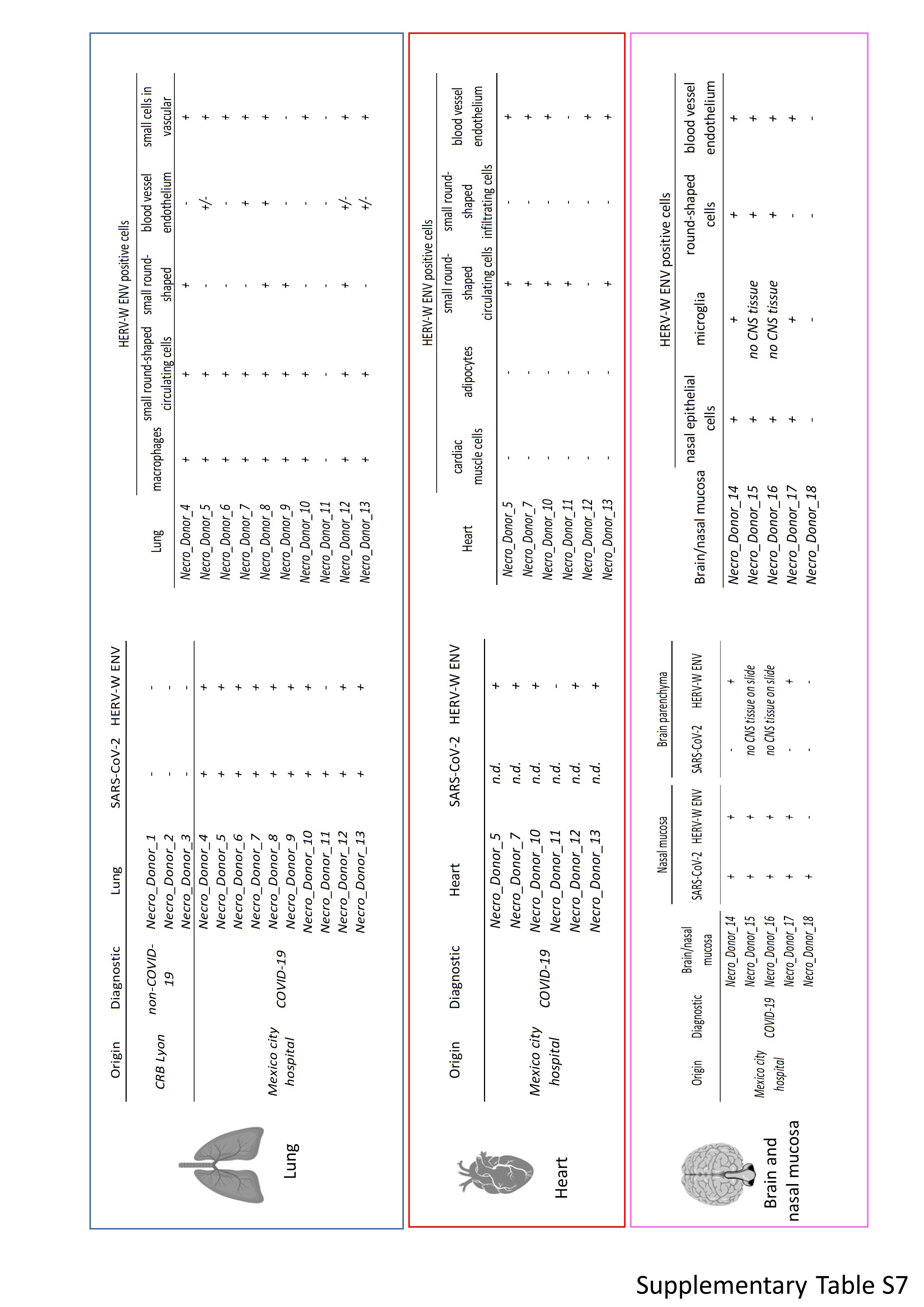
